# Using Test Positivity and Reported Case Rates to Estimate State-Level COVID-19 Prevalence and Seroprevalence in the United States

**DOI:** 10.1101/2020.10.07.20208504

**Authors:** Weihsueh A. Chiu, Martial L. Ndeffo-Mbah

## Abstract

Accurate estimates of infection prevalence and seroprevalence are essential for evaluating and informing public health responses needed to address the ongoing spread of COVID-19 in the United States. A data-driven Bayesian single parameter semi-empirical model was developed and used to evaluate state-level prevalence and seroprevalence of COVID-19 using daily reported cases and test positivity ratios. COVID-19 prevalence is well-approximated by the *geometric mean* of the positivity rate and the reported case rate. As of December 8, 2020, we estimate nation-wide a prevalence of 1.4% [Credible Interval (CrI): 0.8%-1.9%] and a seroprevalence of 11.1% [CrI: 10.1%-12.2%], with state-level prevalence ranging from 0.3% [CrI: 0.2%-0.4%] in Maine to 3.0% [CrI: 1.1%-5.7%] in Pennsylvania, and seroprevalence from 1.4% [CrI: 1.0%-2.0%] in Maine to 22% [CrI: 18%-27%] in New York. The use of this simple and easy-to-communicate model will improve the ability to make public health decisions that effectively respond to the ongoing pandemic.

**Biographical Sketch of Authors:** Dr. Weihsueh A. Chiu, is a professor of environmental health sciences at Texas A&M University. He is an expert in data-driven Bayesian modeling of public health related dynamical systems. Dr. Martial L. Ndeffo-Mbah, is an Assistant Professor of Epidemiology at Texas A&M University. He is an expert in mathematical and computational modeling of infectious diseases.

**Summary Line:** Relying on reported cases and test positivity rates individually can result in incorrect inferences as to the spread of COVID-19, and public health decision-making can be improved by instead using their geometric mean as a measure of COVID-19 prevalence and transmission.

## Introduction

Accurate or reliable estimates of the prevalence and seroprevalence of infection are essential for evaluating and informing public health responses and vaccination strategies to mitigate the ongoing COVID-19 pandemic. The gold standard method to empirically measure disease prevalence and seroprevalence is to conduct periodic large-scale surveillance testing via random sampling (1). However, this approach may be time- and resource-intensive, and only a handful of such surveillance studies has been conducted so far in the United States (US) (2–7). Therefore, public health officials have relied on alternative metrics, such as test positivity, reported cases, fatality rates, hospitalization rates, and epidemiological models’ predictions, to inform COVID-19 responses. Test positivity has, for instance, been commonly used to infer the level of COVID-19 transmission in a population and/or the adequacy of testing (8,9). However, the justifications for use of this metric often reference a WHO recommendation intended to be applied only in a sentinel surveillance context (10), rather than in more general context as it has been frequently implemented. As measures of prevalence, test positivity and reported cases, though readily available and well-understood by public health officials, are very likely to substantially over- and underestimate, respectively, disease transmission/prevalence and seroprevalence (1). Hospitalization and death rates are also similarly readily available, but tend to lag infections by several weeks and only reflect the most severe outcomes (1). Finally, epidemiological models are generally complex mathematical, computational, or statistical models that require extensive data and information for model training, and are perceived as a “black box by most public health practitioners and decision makers (11–13).

Here, we develop a simple semi-empirical model to estimate the prevalence and seroprevalence of COVID-19 at the US state level based only on reported cases, test positivity rate, and testing rate (**Figure 1**). Specifically, we hypothesized that the preferential nature of diagnostic testing for individuals at higher risk of infection in the US is a convex function of the overall testing rate due to the “diminishing return” from expanding general population testing (**Figure 1A-1B**). We modeled this convexity using a negative power function, and calibrated the power parameter using state-wide seroprevalence data, which has only recently become available across all U.S. states (2–7). Infection prevalence estimates were then validated by comparing with two independent, data-driven epidemiological models (11,14,15). We found that the state-level prevalence of COVID-19 in the US can be well-approximated by a weighted average nearly equal to the *geometric mean* of the reported cases and test positivity rates. We evaluated how disease prevalence and seroprevalence varies with changes in reported cases and test positivity, and the implications of applying this simple model on informing public health decision-making to the COVID-19 pandemic in the US. We have implemented our model in an online dashboard (https://wchiu.shinyapps.io/COVID-19-Prevalence-and-Seroprevalence/) to enable easy access by public health officials and the public.

**Figure 1.**
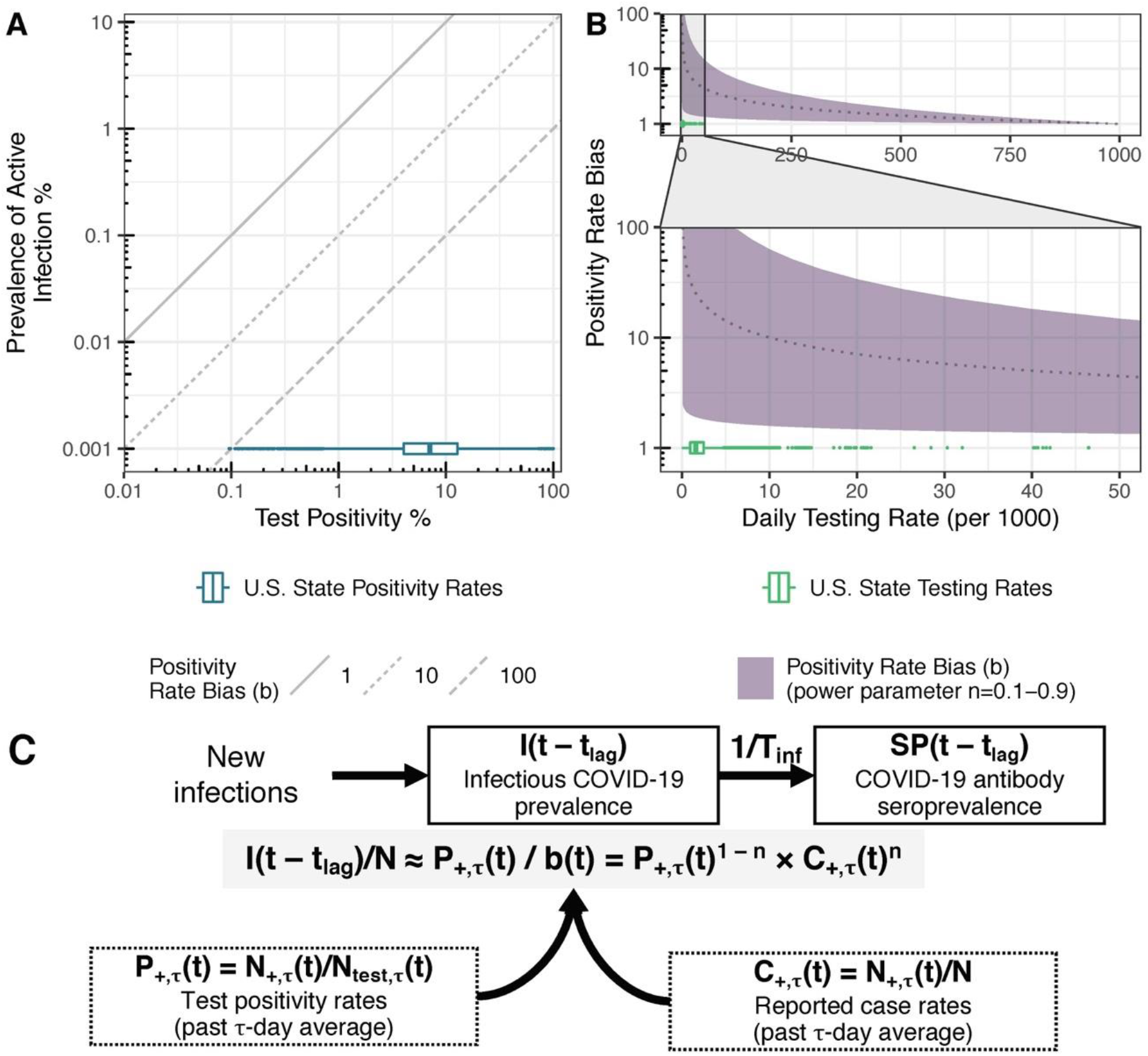
Conceptual model for relationship between test positivity, prevalence of infection, and testing rate. Our model assumes the test positivity rate is correlated to delayed disease prevalence with a bias parameter b(t) modeled as a negative power function of the testing rate b(t) = [N_test,□_(t)/N]^−n^ (equation 2). In **A**), the diagonal lines represent different values of the bias parameter, and the U.S. state positivity rates from March-November are shown for reference. In **B)**, the relationship between testing rate and bias parameter represented by equation (2) is illustrated. Here the shaded region represents different powers n ranging from 0.1 (lower bound bias) to 0.9 (upper bound bias), the dotted line represents n=0.5, and the U.S. state testing rates from March-November are shown for reference. **C)** Compartmental representation of how the relationships between new infections, prevalence I(t), and seroprevalence SP(t) are modeled for each state, given a bias with power n. Observational inputs are the past τ-day averages of number of positive tests N_+,τ_(t) and number of tests performed N_test,τ_(t), the corresponding test positivity rate P_+,τ_(t) and reported case rate C_+,τ_(t), and the state population size N. T_inf_ is the mean infection duration and t_lag_ is the time lag between the last observation t and the time of the inferred prevalence and seroprevalence.

## Methods

### Conceptual basis of a semi-empirical model for the prevalence of COVID-19 infection

Test positivity rate *P*_+,τ_(*t*) = *N*_+,*τ*_(*t*)/*N*_test,*τ*_(*t*) is defined as the percentage of positive diagnostic tests administered over a given period *τ* between *t* − *τ* and *t*. We first hypothesize that, because testing is currently “preferential” (i.e., only those considered more likely to be infected due to symptoms, contacts, etc., are tested), *P*_+,*τ*_(*t*) is correlated to the lagged prevalence *I*(*t* − *t*_lag_)/*N* of COVID-19-infected persons in the population with a time-dependent bias parameter *b*(*t*):

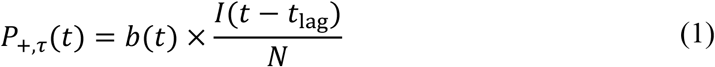

Conceptually, this relationship is shown in **Figure 1A**. We next hypothesize that the bias parameter *b*(*t*) is inversely related to the testing rate *Λ*_*τ*_(*t*) = *N*_test,*τ*_(*t*)/*N* over the same period *τ*. Clearly, at a testing rate of 1, where everyone is tested, there is no bias, so *b* = 1. On the other hand, for very low testing rates, the bias is likely to be high, as only the most severely ill will be tested. For instance, if all COVID-19 deaths are tested daily, but no one else, then the positivity rate will be equal to 100% and the bias will be equal to the inverse of the infection prevalence, *b* = *N*/*I*(*t* − *t*_lag_). Moreover, at low testing rates, increasing the testing rate will likely preferentially increase the infected population testing rate relative to the general population testing rate, so *b*(*t*) will decline more rapidly than at higher testing rates, as there is “diminishing return” from increased testing. Thus, it is reasonable to assume that *b*(*t*) is a convex function of *Λ*_*τ*_(*t*). We therefore model the bias as a negative power function of *Λ*_*τ*_(*t*):

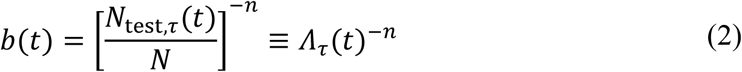

While this appears to imply an unbounded bias as the testing rate goes to zero, as shown below, our model will naturally limit the bias parameter when test positivity is 100%. Combining equations (1)-(2), and re-arranging leads to the following relationship between test positivity and the infectious population:

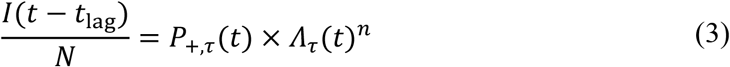

Additionally, because test positivity and the testing rate share a term *N*_test,*τ*_(*t*), equation (3) can be further rearranged as

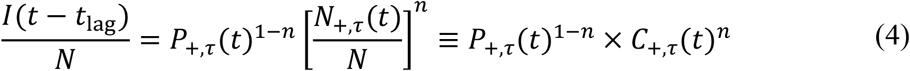

where the last term is the reported cases per capita *C*_+,*τ*_(*t*) = *N*_+,*τ*_(*t*)/*N*. Thus, our hypothesis predicts that the infectious population is proportional to a *weighted geometric mean* of the positivity rate and the reported case rate, with *n* = ½ corresponding to equal weighting.

We can also rearrange equation (1) and view the bias parameter as the relative efficacy of testing infected individuals compared to the general population:

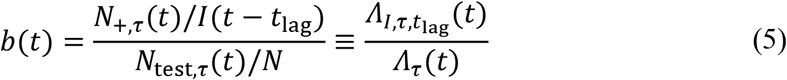

Here,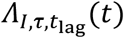 is the daily rate of testing of infectious individuals (with averaging time *τ* and lag *t*_lag_), whereas *Λ*_*τ*_(*t*) is the daily rate of testing of the general population, as previously defined. Thus, the bias reflects the extent to which infectious individuals are “preferentially” tested. Moreover, due to the way *I*(*t* − *t*_lag_) is calculated, when positivity is 100% so that *N*_+,*τ*_(*t*) = *N*_test,*τ*_(*t*), the bias appropriately equals *N*/*I*(*t* − *t*_lag_).

We use this semi-empirical model for infection prevalence to estimate seroprevalence SP(*t*) using a simple first order, one-compartment model that depends only on the mean infection duration *T*_inf_ (**Figure 1C**). Because testing data are available on a daily basis, we use a discrete time model SP(*t*) = SP(*t* − 1) + *I*(*t* − 1)/*T*_inf_. The seroprevalence at any time *t* is then the cumulative sum of the infection prevalence values at times < *t* divided by the infection duration 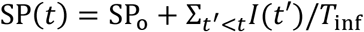. As an initial condition, we allow for an offset SP_o_ for missed infections during the early part of the pandemic before regular and large-scale testing was established. Therefore, combining with equation (4) gives the seroprevalence as:

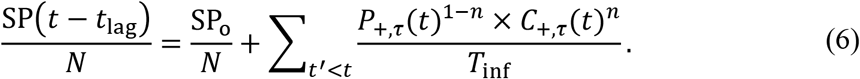

Equations (4) and (6) therefore comprise the complete semi-empirical model for infection prevalence and seroprevalence based solely on positivity *P*_+,*τ*_(*t*) and the reported case rates *C*_+,*τ*_(*t*), which we calculate from data obtained from the COVID Tracking Project (16), averaged of the past *τ* days. We fix the averaging time *τ* at 14 days, and the lag time *t*_lag_ at 7 days, so the semi-empirical model has only three remaining free parameters: the power parameter *n*, the infection during *T*_inf_, and the initial condition for seroprevalence SP_o_.

### Bayesian calibration to seroprevalence data

To calibrate the model, we utilized state-wide seroprevalence data, which has only recently become available for all 50 states and the District of Columbia (**Table S1**). We used a Bayesian MCMC approach to calibrate the model to the empirical data (Table 1) and the potential scale reduction factor (PSRF) was used to assess convergence, with a value of <1.2 regarded as adequate (17,18). Details about model calibration are found in the Supplementary Methods.

**Table 1.** Model parameters, prior and posterior distributions, and convergence diagnostic

| Model<br>parameter | Prior distributions | Prior<br>Source | Posterior median<br>[95% CrI] | PSRF |
| --- | --- | --- | --- | --- |
| $n$<br>(power parameter) | $\mu$: Uniform on $\text{logit}(n)$<br>$\Sigma$: Log-Uniform | Non-<br>informative<br>prior | $\mu$: 0.47 [0.41 - 0.57]<br>$\Sigma$: 0.20 [0.11 – 0.32] | $\mu$: 1.08<br>$\Sigma$: 1.00 |
| $SP_0$<br>(initial condition<br>for<br>seroprevalence) | $\mu$: Uniform on<br>$\text{logit}(SP_0)$<br>$\Sigma$: Log-Uniform | Non-<br>informative<br>prior | $\mu$: 0.46% [0.03% -<br>1.26%]<br>$\Sigma$: 1.48 [0.91 – 3.19] | $\mu$: 1.03<br>$\Sigma$: 1.01 |
| $T_{\text{inf}}$<br>(infection<br>duration in days) | $\mu$:<br>Normal($m=14, sd=3.5$) | (25) | $\mu$: 15.2 [8.2 – 21.4] | $\mu$: 1.06 |
| $\sigma_{\text{err}}$<br>(residual standard<br>error on natural<br>log scale) | $\mu$: Log-Uniform | Non-<br>informative<br>prior | $\mu$: 0.28 [0.23 – 0.33] | $\mu$: 1.00 |

$\mu$ =fixed effect, $\Sigma$ = random effect standard deviation (on logit scale), PSRF=potential scale reduction factor convergence diagnostic (17).

### Validation of prevalence estimates with epidemiological models

For validation, we compare prevalence estimates to those of two models of SARS-CoV-2 transmission in U.S. states are considered here: a Bayesian extended-SEIR model (15) (run through July 22, 2020) and a Bayesian semi-mechanistic model by Imperial College (19) (run through July 20, 2020), which applies the model published by Flaxman et al., 2020 (11) to the US. These models are calibrated at the US state-level to reported deaths and/or cases, but do not utilize test positivity or testing rate as an input. Both of these are Bayesian models, and we use these models’ posterior distributions for comparison.

### Bias of test positivity and reported cases in estimating prevalence and seroprevalence

Our model can be used to estimate the degree of bias in current measures of prevalence (test positivity and reported case rates) and seroprevalence (cumulative reported cases). The over-reporting bias of test positivity as a measure of prevalence is already given in equation (2). The under-reporting bias of reported case rates can be calculated by rearranging equation (4),

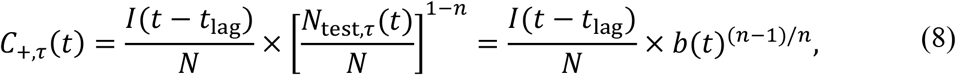

so the under-reporting bias is *b*(*t*)^(*n*− 1)/*n*^, which is equal to *b*(*t*) for *n* = ½. The implied bias from cumulative reported cases as a measure of seroprevalence is calculated by dividing by sum of *C*_+,*τ*_(*t*) by the seroprevalence estimated by equation (6).

### Software

All analyses were performed using the R statistical software (R version 3.6.1) in RStudio (Version 1.2.1335). We developed a publicly available online dashboard, using R and Shiny, to analyze and visualize state-level estimates of COVID-19 prevalence and seroprevalence from our semi-empirical model (https://wchiu.shinyapps.io/COVID-19-Prevalence-and-Seroprevalence/).

## Results

### Bayesian calibration to seroprevalence data

Four independent Markov chain Monte Carlo chains were simulated, and reached adequate convergence after 20,000 iterations per chain, with PSRF < 1.1 for all parameters (see **Table 1** and **Table S2**) and the multivariate PSRF=1.07. For inference, 2,000 samples were selected randomly from across the 80,000 available iterations.

The 95% credible intervals for the power parameter *n* include 0.5 (corresponding to an unweighted geometric mean) both for the fixed effect as well as for the state-specific random effects (**Table S2, Figure S1**). For the seroprevalence offset SP_o_, the posterior median for most states was < 1% initial condition, but two states had posterior medians > 5%. For NY, it was estimated that 14% [95% CrI: 7.5%-20%] of initial cases were missed, and for LA, the posterior estimate was 5.0% [CrI: 1.6%-8.6%]. These values are consistent with these two states having large initial surges when testing was highly limited, and therefore were likely to have missed a large number of cases. The large variation in SP_o_ values across states is consistent with high heterogeneity that has been noted both in the size of their initial surge of infections and in their testing capacity and availability.

Comparison of posterior estimates and observations by state are shown in **Figure 2**, and show the model to be highly consistent with available seroprevalence data both in terms of level and trends. The residual error was estimated to have a standard deviation of 0.28 [CrI: 0.23-0.33] on the natural log scale, corresponding to a coefficient of variation of 29% [CrI: 23%-33%], and the R^2^ between the posterior median and the observed point estimates was 0.76 (**Figure S2**).

**Figure 2.**
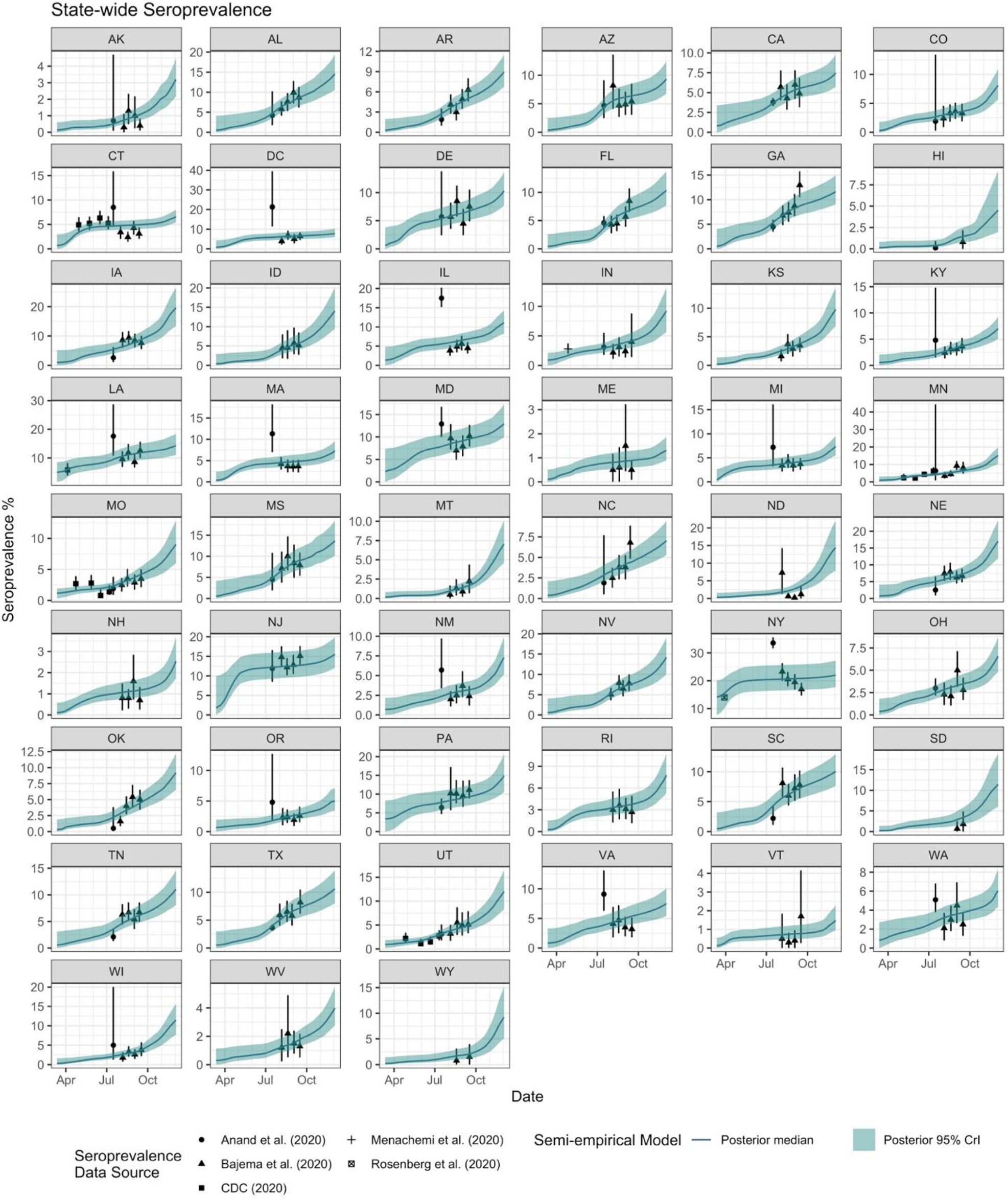
Calibration results of our semi-empirical model for COVID-19 antibody seroprevalence (posterior median and 95% credible intervals) for each state with state-wide seroprevalence data (reported point estimates and 95% confidence intervals shown).

### Validation of prevalence estimates with epidemiological models

We next compared our model estimates for the prevalence of active infections with those from two independent epidemiologic models of U.S. states. As shown in **Figure 3**, the posterior estimates of the semi-empirical model are highly consistent with those from the epidemiologic models. The residual standard error (RSE) difference between the posterior medians of semi-empirical estimate and the extended SEIR model is 0.52 natural log units (see **Figure S3**), corresponding to a coefficient of variation of 56%, and have an R^2^ of 0.75. Similarly, the comparison with the Imperial model yields an RSE of 0.73, corresponding to an 84% coefficient of variation, and an R^2^ of 0.72. These RSE values should be taken in context of the posterior uncertainty in the epidemiologic models themselves, which have individual uncertainties corresponding to coefficients of variation of 45% and 23% for the extended SEIR and Imperial models, respectively, as well as the differences between the two models, which have a coefficient of variation of 63%. Overall, the semi-empirical estimate of infection prevalence is consistent with the results of the available epidemiologic models.

**Figure 3.**
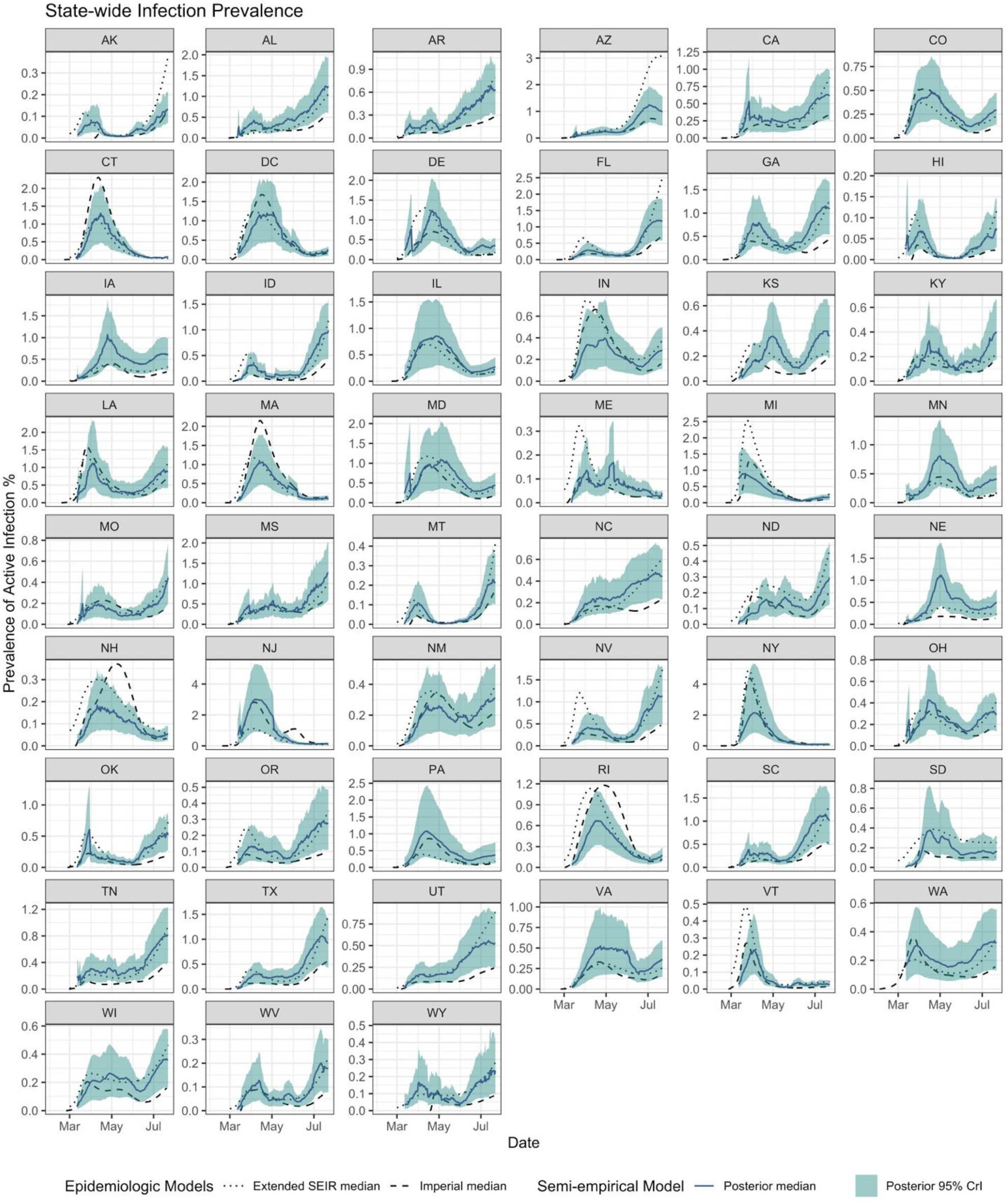
Validation of COVID-19 infection prevalence estimates (posterior median and 95% credible interval) for each state in comparison to posterior median estimates from two data-driven epidemiologic models: an extended-SEIR model calibrated to reported cases and confirmed deaths through July 22, 2020 (15) and a semi-mechanistic model calibrated to confirmed deaths through July 20, 2020 by Imperial College (19)).

### Current estimates of prevalence and seroprevalence

As of December 8, 2020, our calibrated and validated semi-empirical model estimates that in the US, infection prevalence was 1.4% [CrI: 0.8%-1.9%] with a seroprevalence of 11.1% [CrI: 10.1%-12.2%] (**Figures 4, S4; Table S3**). In individual states, estimated prevalence ranged from 0.3% [CrI: 0.2%-0.4%] in Maine to 3.0% [CrI: 1.1%-5.7%] in Pennsylvania, with 8 states having at least 2% prevalence, and test positivity having a bias *b* between five and fifteen (**Figure S4A**). The two-week trend in estimated prevalence was increasing in all but 15 states (**Figure 4A**). Estimated seroprevalences in individual states ranged from 1.4% [CrI: 1.0%-2.0%] in Maine to 22% [CrI: 18%-27%] in New York, with 9 states exceeding 15%, and cumulative reported cases typically accounting for around one in three of estimated total cases (**Figures 4B, S4B**).

**Figure 4.**
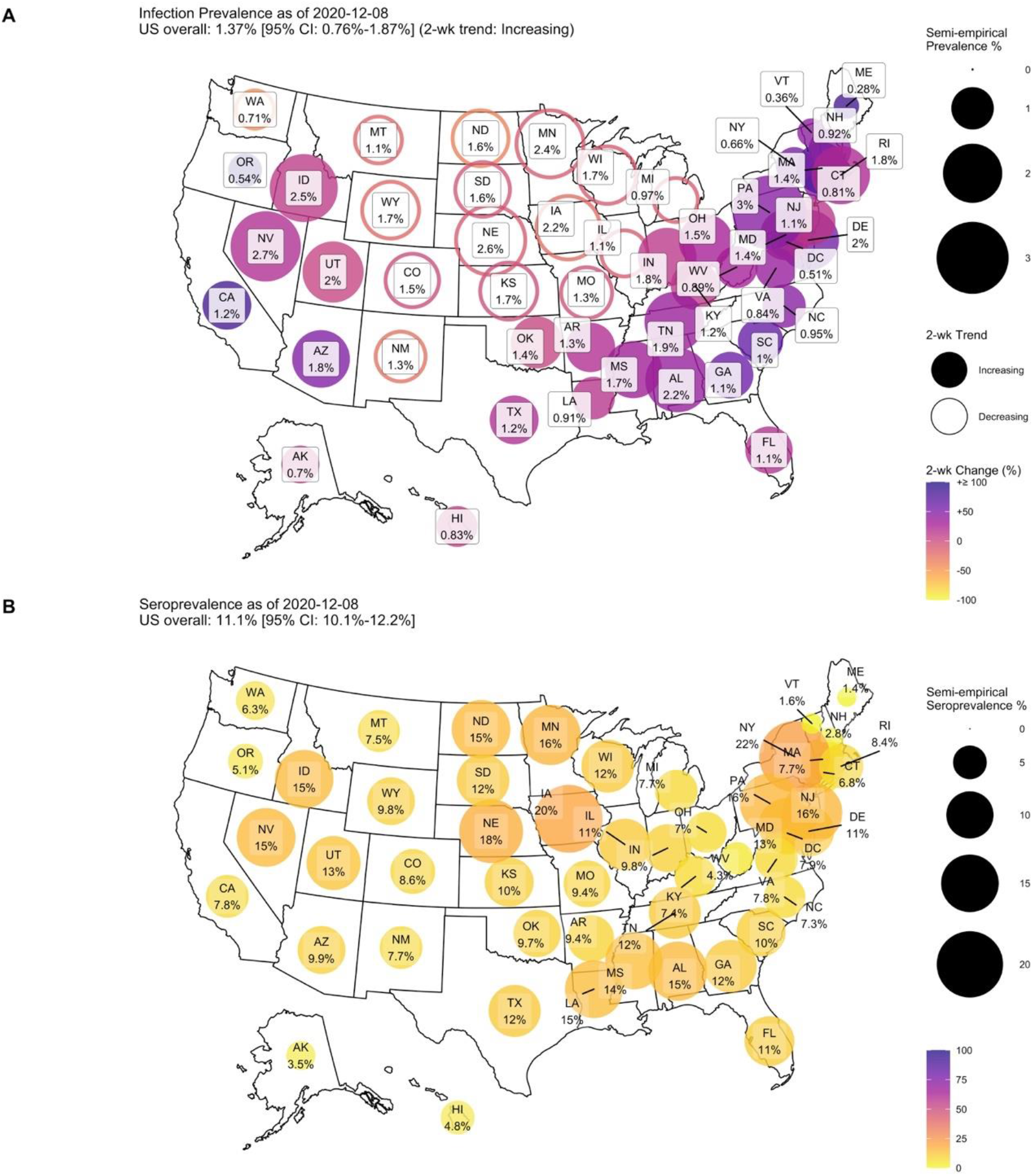
**A)** Map of estimated prevalence and transmission trends as of December 8, 2020, based on data through December 15, 2020. **B)** Map of estimated seroprevalence as of December 8, 2020, based on data through December 15, 2020.

Between April 1 and December 8, 2020, the test positivity rate bias *b* and the ratio between estimated seroprevalence and cumulative reported cases were shown to decrease over time (**Figure S5**). In April, the median positivity rate bias across states was 36 and the cumulative cases underreporting bias ranged between 0.01 to 0.12 (i.e., only 1%-12% of cases were reported). By the beginning of December, the median positivity rate bias had declined to 13 and the cumulative cases underreporting bias ranged between 0.14 and 1 (**Figures S5A, S5C**). Across the U.S. in aggregate, from April to December, the median positivity rate bias declined from 66 to 10, and the cumulative cases underreporting bias improved from 0.03 to 0.33 (**Figures S5B, S5D**).

The pitfalls of relying on reported cases or test positivity rate alone to estimate the course of the epidemic are illustrated for three states, MN, VA, and WI, where reported cases and positivity trends were in opposite directions in May and/or August (**Figure S6**). Specifically, in May, reported cases were rising substantially in MN, VA, and WI at the same time that the test positivity rate was declining, testing rate was increasing, and the model predicted prevalence was flat or decreasing (**Figure S6**). By contrast, in August, the states of MN and WI all showed declining reported case rates while positivity was increasing, while our model predicts that COVID-19 prevalence was actually increasing during this time. In both scenarios, the increase (decrease) in reported cases was due to expanded (declining) testing rates, respectively.

## Discussion

Reported case rates and test positivity rates have been widely used to inform or justify public health decisions, such as increasing or relaxing non-pharmaceutical interventions, for the control of the COVID-19 pandemic in the US (20,21). A recent report of the National Academies of Sciences and Engineering Medicine (NASEM) has urged caution about the reliability/validity of directly using data such as reported case rates and test positivity rates to inform decision making for COVID-19 (1). Though these data are usually readily available, the NASEM report concludes that they are likely to substantially underestimate or overestimate the real state of disease spread (1). Therefore, there is a critical need to develop simple and more reliable data-driven metrics/approaches to inform local public health decision-making.

We have developed a simple semi-empirical approach to estimate the prevalence and seroprevalence of COVID-19 infections in a population using only reported cases and testing rates that does not require developing and maintaining a complex, data-driven mathematical model. Based on a simple hypothesis that the bias in test positivity is a convex, negative power function of the testing rate, we find that the COVID-19 prevalence, with a 1-week lag, is well-approximated by the *geometric mean* of the positivity rate and the reported case rate averaged over the last 2 weeks. Seroprevalence can then be calculated in a straight-forward manner by taking a cumulative sum while accounting for the duration between infection and seropositivity, a period of typically 2 weeks, and a state-specific offset accounting for missed infections in the early part of the pandemic prior to establishment of regular testing (equation 6). Our model resulted in an accurate fit to recently available state-level seroprevalence data from across the U.S. Additionally, the prevalence estimates of our semi-empirical model were shown to compare favorably to those from two data-driven epidemiological models. We estimate nation-wide prevalence rate as of December 8, 2020 to be 1.4% [CrI: 0.8%-1.9%], corresponding to a test positivity bias of around 10, and nation-wide seroprevalence to be 11.1% [CrI: 10.1%-12.2%], so that cumulative reported cases correspond to approximately one-third of actual infections. At the state level, estimated seroprevalence was one to seven times cumulative reported cases. These estimates compare favorably to those previously published using more complicated approaches (22,23).

Our analysis suggests that public health policy related to either non-pharmaceutical (masking and social distancing) or pharmaceutical interventions (vaccination) may be informed by available data in four main ways:

- First, prevalence should be inferred as decreasing only if both reported cases and positivity rates are continuously declining without decreasing testing rates. This criterion can be taken as a surrogate for *R*_*eff*_(*t*) < 1, since declining prevalence is a sufficient condition for an effective reproduction number below one. When there is a divergence in direction of the trends between reported cases and positivity (**Figure S6**), the trend in their geometric mean can be used as an indicator of increasing or decreasing prevalence.
- Second, the gap between reported case and test positivity rates should be decreasing in order to ensure improvement in the sufficiency of testing. For most of the U.S., this gap has hovered between 100- and 1000-fold. For example, in MN, VA, and WI (**Figure S6**), as testing ramped up in May, the gap narrowed; however, the gap widened in August, indicating that testing rates needed to be brought back up (24). Indeed, as noted by the NASEM report, absolute testing rates, irrespective of positivity rates, may be a better indicator of the sufficiency of testing.
- Third, reported cases, test positivity, and testing rates should be publicly reported at the county or municipal level in order to provide local governments, health agencies, medical personnel, and the public with the necessary information to evaluate local pandemic conditions. Currently, only reported cases are routinely provided at the local level, with positivity and testing rates comprehensively aggregated only at the state level.
- Finally, seroprevalence estimates can play a key role in forecasting future potential spread of the pandemic and threshold vaccination coverage needed to stamp out disease transmission at the state or community-level.

As with any model, ours has a number of limitations. The most significant limitation is the lack of more comprehensive, random sampling-based data with which to further validate the model. However, our model did accurately fit all the available seroprevalence data, including recent CDC data at multiple time points across all 50 states and the District of Columbia (6). As further validation of the approach, we applied our model internationally to 15 countries for which both nation-wide seroprevalence data (**Table S4**) and daily testing data were available. The 95% CrI for our model, using the random effects posterior distributions from our U.S. state-level calibration, covered all the seroprevalence data except for Russia (**Figure S7**), suggesting that this approach might be more broadly applicable, though requiring nation-specific calibration. With respect to prevalence, we could only compare to epidemiologic model-based estimates of prevalence due to lack of random sample-based surveillance data. However, we believe this limitation is mitigated by our use of two independent estimates with completely different model structures, one of which is a more traditional extended-SEIR model, and other of which is a “semi-mechanistic” model partially statistical in nature. Another important limitation is the relatively limited range of testing rate observations for most U.S. states (**Figure 1B**). For this reason, we cannot necessarily guarantee that our results can be easily extrapolated to substantially higher testing rates. However, with higher testing rates, the difference between test positivity rates and reported case rates would decrease and reduce the effect of greater uncertainty in the degree of bias between the test positivity rate and the lagged prevalence. Finally, we did not address the sensitivity and specificity of diagnostic testing; however, the impact of imperfect test accuracy is likely to have a minimal impact on our results.

In conclusion, we found that the COVID-19 prevalence is well-approximated by the *geometric mean* of the positivity rate and the reported case rate, and that seroprevalence can be estimated by taking a cumulative sum while accounting for the duration between infection and seropositivity, a period of typically 2 weeks, and a state-specific offset. The use of this simple, reliable, and easy-to-communicate approach to estimating COVID-19 prevalence and seroprevalence will improve the ability to make public health decisions that effectively respond to the ongoing COVID-19 pandemic in the U.S.

## Data Availability

The codes and data used to generate our results are available on GitHub

https://github.com/wachiuphd/COVID-19-US-Semi-Empirical

## Acknowledgement

We thank the COVID Tracking Project and Our World in Data for compiling COVID-19 case and testing data and providing it to the public.

