## Supplemental Materials for "Using Test Positivity and Reported Case Rates to Estimate State-Level COVID-19 Prevalence and Seroprevalence in the United States"

**Supplemental Methods**

***Bayesian calibration to seroprevalence data***

To calibrate the model, we utilized state-wide seroprevalence data, which has only recently become available for all 50 states and the District of Columbia (**Table S1**). We only included data from states where the entire state was sampled or where adjustments for convenience sampling bias were integrated into the analysis (1–6), and verified our selections by comparison to three independent systematic reviews (7–9). Data were extracted from study tables or supplement materials. For 4 states (CT, LA, MO, UT), additional data were downloaded from the CDC Dashboard for Commercial Laboratory Surveys (10).

We used a Bayesian approach to estimate model parameters, using the prior distributions shown in **Table 1**. Because states differ somewhat in their testing availability and capacity, as well as in the extent to which the initial surge of infections was missed, we assume random effects for the power parameter *n* and the initial condition for seroprevalence SP_o_. We use a logit transformation to restrict values between 0 and 1, where SP_o_ is scaled by state population, and assume state-to-state variation on the transformed values is distributed normally with unknown random effects standard deviation Σ. We use non-informative prior distributions consisting of a uniform distribution on the logit scale for the fixed effects μ, and a log-uniform distribution for Σ. For the duration of infection *T*_inf_, we assign an informative prior distribution with mean 14 days and standard deviation 3.5 days, corresponding to a 95% CI range of 7-21 days, based on the CDC estimate that it takes 7-21 days after infection to become seropositive (11).

The likelihood function assumes independent log-normal distributed errors given an observed seroprevalence $y_{\mathrm{obs}}$ and estimated seroprevalence (equation 6) $y_{\mathrm{pred}}$:

$$\ln y_{\mathrm{obs}}=\ln y_{\mathrm{pred}}+\epsilon$$

$$\epsilon\sim N(0,\sigma_{\mathrm{obs}}^{2}+\sigma_{\mathrm{err}}^{2})$$

(7)

with the observed error variance $\sigma_{\mathrm{obs}}^{2}$ estimated from reported 95% CI for each observation $[\sigma_{\mathrm{obs}}=(\ln{UCL}_{\mathrm{obs}}-\ln y_{\mathrm{obs}})/z_{97.5}]$, where ${UCL}_{\mathrm{obs}}$ is the reported upper confidence limit and $z_{97.5}$ is the z-score of the 97.5^th^ percentile (in some cases, the reported lower confidence limit was 0, so only the upper confidence limit was used to calculate $\sigma_{\mathrm{obs}}^{2}$). The residual error standard deviation $\sigma_{\mathrm{err}}$ is given a log-uniform prior.

Markov chain Monte Carlo (MCMC) simulation was conducted to sample from the posterior distribution. Sampling was conducted using component-wise Metropolis sampling with a normal proposal distribution (12). The variance of the proposal distribution was adjusted for each parameter to maintain an acceptance rate between 20% and 60%. Four independent chains with different starting points and random seeds were run, and the potential scale reduction factor (PSRF) was used to assess convergence, with a value of <1.2 regarded as adequate (13,14).

***Data***

U.S. testing and reported case data were downloaded from the COVID Tracking Project using their API (15). In the data definitions, the fields “positiveIncrease” (confirmed plus probable [if reported] cases of COVID-19 reported) and “negativeIncrease” (increase in unique people with completed PCR test that returns negative) were used to represent daily reported cases and negative test results, respectively. International testing and case data were downloaded from Our World in Data (16) or the U.K. government (17).

**Supplemental Tables and Figures**

**Table S1.** State-wide seroprevalence data

| **State** | **Min date** | **Max date** | **Point est. (%)** | **LCL (%)** | **UCL (%)** | **Source** |
| --- | --- | --- | --- | --- | --- | --- |
| NY | 3/29/20 | 3/29/20 | 14 | 13.3 | 14.7 | (3) |
| IN | 4/25/20 | 4/29/20 | 2.79 | 2.02 | 3.7 | (2) |
| CT | 4/26/20 | 5/3/20 | 4.9 | 3.6 | 6.5 | (1,6) |
| CT | 5/21/20 | 5/26/20 | 5.2 | 3.9 | 6.6 | (1,6) |
| CT | 6/15/20 | 6/17/20 | 6.3 | 5 | 7.8 | (1,6) |
| CT | 7/3/20 | 7/6/20 | 5.2 | 4 | 6.7 | (1,6) |
| LA | 4/1/20 | 4/8/20 | 5.8 | 3.9 | 8.2 | (1,6) |
| MN | 4/30/20 | 5/12/20 | 2.4 | 1 | 4.5 | (1,6) |
| MN | 5/25/20 | 6/7/20 | 2.2 | 1.4 | 3.3 | (1,6) |
| MN | 6/15/20 | 6/27/20 | 4.3 | 3.4 | 5.8 | (1,6) |
| MN | 7/6/20 | 7/17/20 | 6.1 | 4.4 | 8.3 | (1,6) |
| MO | 4/20/20 | 4/26/20 | 2.7 | 1.7 | 3.9 | (1,6) |
| MO | 5/25/20 | 5/30/20 | 2.8 | 1.7 | 4.1 | (1,6) |
| MO | 6/15/20 | 6/20/20 | 0.8 | 0.6 | 1.8 | (1,6) |
| MO | 7/5/20 | 7/9/20 | 1.4 | 0.9 | 2.5 | (1,6) |
| UT | 4/20/20 | 5/3/20 | 2.2 | 1.2 | 3.4 | (1,6) |
| UT | 5/25/20 | 6/5/20 | 1.1 | 0.6 | 2.1 | (1,6) |
| UT | 6/15/20 | 6/27/20 | 1.5 | 0.9 | 2.6 | (1,6) |
| UT | 7/6/20 | 7/15/20 | 2.7 | 1.8 | 3.9 | (1,6) |
| AK | 7/1/20 | 7/31/20 | 0.7 | 0.1 | 4.7 | (4) |
| AL | 7/1/20 | 7/31/20 | 4.2 | 1.8 | 10.2 | (4) |
| AR | 7/1/20 | 7/31/20 | 1.9 | 1 | 3.5 | (4) |
| AZ | 7/1/20 | 7/31/20 | 4.7 | 2.5 | 9.1 | (4) |
| CA | 7/1/20 | 7/31/20 | 3.8 | 3.3 | 4.3 | (4) |
| CO | 7/1/20 | 7/31/20 | 1.9 | 0.3 | 13.4 | (4) |
| CT | 7/1/20 | 7/31/20 | 8.5 | 4.6 | 15.9 | (4) |
| DC | 7/1/20 | 7/31/20 | 21.3 | 11.4 | 39.5 | (4) |
| DE | 7/1/20 | 7/31/20 | 5.7 | 2.4 | 13.8 | (4) |
| FL | 7/1/20 | 7/31/20 | 4.6 | 3.6 | 5.8 | (4) |
| GA | 7/1/20 | 7/31/20 | 4.5 | 3.5 | 5.9 | (4) |
| HI | 7/1/20 | 7/31/20 | 0.1 | 0 | 0.9 | (4) |
| IA | 7/1/20 | 7/31/20 | 2.6 | 1.1 | 6.2 | (4) |
| IL | 7/1/20 | 7/31/20 | 17.5 | 15.2 | 20.2 | (4) |
| IN | 7/1/20 | 7/31/20 | 3.1 | 1.8 | 5.5 | (4) |
| KY | 7/1/20 | 7/31/20 | 4.8 | 1.5 | 14.8 | (4) |
| LA | 7/1/20 | 7/31/20 | 17.6 | 10.8 | 28.7 | (4) |
| MA | 7/1/20 | 7/31/20 | 11.3 | 7 | 18.2 | (4) |
| MD | 7/1/20 | 7/31/20 | 12.9 | 10 | 16.7 | (4) |
| MI | 7/1/20 | 7/31/20 | 7.2 | 3.2 | 16.1 | (4) |
| MN | 7/1/20 | 7/31/20 | 6.3 | 0.9 | 44.4 | (4) |
| MO | 7/1/20 | 7/31/20 | 1.9 | 0.9 | 3.8 | (4) |
| MS | 7/1/20 | 7/31/20 | 4.5 | 1.9 | 10.8 | (4) |
| NC | 7/1/20 | 7/31/20 | 1.9 | 0.5 | 7.7 | (4) |
| NE | 7/1/20 | 7/31/20 | 2.5 | 0.9 | 6.5 | (4) |
| NJ | 7/1/20 | 7/31/20 | 11.9 | 8.5 | 16.6 | (4) |
| NM | 7/1/20 | 7/31/20 | 5.7 | 3.4 | 9.7 | (4) |
| NY | 7/1/20 | 7/31/20 | 33.6 | 31.7 | 35.6 | (4) |
| OH | 7/1/20 | 7/31/20 | 3 | 2.2 | 4.1 | (4) |
| OK | 7/1/20 | 7/31/20 | 0.5 | 0.1 | 3.8 | (4) |
| OR | 7/1/20 | 7/31/20 | 4.8 | 1.8 | 12.7 | (4) |
| PA | 7/1/20 | 7/31/20 | 6.4 | 4.7 | 8.8 | (4) |
| SC | 7/1/20 | 7/31/20 | 2.2 | 1.1 | 4.4 | (4) |
| TN | 7/1/20 | 7/31/20 | 2.1 | 1.3 | 3.6 | (4) |
| TX | 7/1/20 | 7/31/20 | 3.6 | 3.1 | 4.2 | (4) |
| UT | 7/1/20 | 7/31/20 | 3.1 | 1.8 | 5.1 | (4) |
| VA | 7/1/20 | 7/31/20 | 9.1 | 6.3 | 13.1 | (4) |
| WA | 7/1/20 | 7/31/20 | 5.1 | 3.8 | 6.8 | (4) |
| WI | 7/1/20 | 7/31/20 | 5 | 1.3 | 20 | (4) |
| AK | 8/6/20 | 8/11/20 | 0.3 | 0 | 1.12 | (5) |
| AL | 7/29/20 | 8/13/20 | 5.8 | 4.16 | 7.71 | (5) |
| AR | 7/29/20 | 8/13/20 | 4.1 | 2.74 | 5.63 | (5) |
| AZ | 7/31/20 | 8/11/20 | 8.2 | 4.11 | 13.59 | (5) |
| CA | 7/30/20 | 8/5/20 | 5.7 | 4.05 | 7.77 | (5) |
| CO | 7/30/20 | 8/7/20 | 2.4 | 0.89 | 4.53 | (5) |
| CT | 7/30/20 | 8/3/20 | 3.4 | 2.04 | 4.72 | (5) |
| DC | 7/30/20 | 8/13/20 | 3.9 | 2.18 | 5.76 | (5) |
| DE | 7/29/20 | 8/13/20 | 5.7 | 3.55 | 8.22 | (5) |
| FL | 7/31/20 | 8/3/20 | 4.3 | 2.77 | 5.86 | (5) |
| GA | 8/2/20 | 8/11/20 | 6.8 | 4.83 | 8.81 | (5) |
| HI | 8/3/20 | 8/11/20 |  |  |  | (5) |
| IA | 7/29/20 | 8/13/20 | 8.6 | 5.98 | 11.34 | (5) |
| ID | 8/4/20 | 8/11/20 | 4.5 | 1.8 | 7.99 | (5) |
| IL | 7/29/20 | 8/10/20 | 3.9 | 2.52 | 5.21 | (5) |
| IN | 7/31/20 | 8/11/20 | 2.2 | 1.1 | 3.63 | (5) |
| KS | 7/29/20 | 8/8/20 | 1.6 | 0.7 | 2.8 | (5) |
| KY | 7/30/20 | 8/13/20 | 2.4 | 1.32 | 3.62 | (5) |
| LA | 7/28/20 | 8/13/20 | 9.6 | 6.88 | 12.29 | (5) |
| MA | 7/30/20 | 8/10/20 | 4.2 | 2.89 | 5.81 | (5) |
| MD | 7/31/20 | 8/11/20 | 9.7 | 7.26 | 12.87 | (5) |
| ME | 7/30/20 | 8/11/20 | 0.5 | 0 | 1.16 | (5) |
| MI | 7/30/20 | 8/11/20 | 3.4 | 2.16 | 4.84 | (5) |
| MN | 7/29/20 | 8/13/20 | 3.5 | 2.06 | 5.01 | (5) |
| MO | 7/28/20 | 8/10/20 | 2.5 | 1.44 | 3.61 | (5) |
| MS | 7/30/20 | 8/13/20 | 7.1 | 3.75 | 11.09 | (5) |
| MT | 7/29/20 | 8/10/20 | 0.5 | 0 | 1.67 | (5) |
| NC | 7/29/20 | 8/10/20 | 2.5 | 1.31 | 3.72 | (5) |
| ND | 7/29/20 | 8/12/20 | 7.3 | 1.3 | 14.28 | (5) |
| NE | 7/28/20 | 8/13/20 | 7.4 | 5.25 | 9.75 | (5) |
| NH | 7/30/20 | 8/11/20 | 0.8 | 0.22 | 1.48 | (5) |
| NJ | 7/31/20 | 8/11/20 | 14.8 | 12.23 | 17.57 | (5) |
| NM | 7/29/20 | 8/13/20 | 2 | 1.09 | 3.02 | (5) |
| NV | 7/30/20 | 8/2/20 | 5.1 | 3.63 | 6.66 | (5) |
| NY | 7/31/20 | 8/11/20 | 23.3 | 20.07 | 26.32 | (5) |
| OH | 7/29/20 | 8/11/20 | 2.3 | 1.11 | 3.61 | (5) |
| OK | 7/28/20 | 8/4/20 | 1.6 | 0.85 | 2.49 | (5) |
| OR | 8/5/20 | 8/11/20 | 2.3 | 1.03 | 3.84 | (5) |
| PA | 7/31/20 | 8/11/20 | 10.2 | 5.71 | 17.21 | (5) |
| PR | 7/27/20 | 8/7/20 | 1.1 | 0.43 | 1.8 | (5) |
| RI | 7/30/20 | 8/11/20 | 3 | 1.23 | 5.51 | (5) |
| SC | 7/30/20 | 8/13/20 | 8.1 | 5.79 | 10.69 | (5) |
| SD | 7/29/20 | 8/12/20 |  |  |  | (5) |
| TN | 7/30/20 | 8/11/20 | 6.3 | 4.41 | 8.25 | (5) |
| TX | 7/29/20 | 8/5/20 | 5.9 | 4.04 | 7.95 | (5) |
| UT | 7/30/20 | 8/11/20 | 3.2 | 1.7 | 5.03 | (5) |
| VA | 7/31/20 | 8/11/20 | 4.1 | 1.8 | 6.93 | (5) |
| VT | 7/30/20 | 8/11/20 | 0.5 | 0 | 1.84 | (5) |
| WA | 7/29/20 | 8/11/20 | 2.1 | 0.81 | 3.7 | (5) |
| WI | 7/30/20 | 8/13/20 | 1.8 | 0.76 | 3.18 | (5) |
| WV | 7/30/20 | 8/13/20 | 1.2 | 0.23 | 2.5 | (5) |
| WY | 7/29/20 | 8/11/20 |  |  |  | (5) |
| AK | 8/12/20 | 8/26/20 | 1.3 | 0.5 | 2.33 | (5) |
| AL | 8/12/20 | 8/26/20 | 7.6 | 5.33 | 9.76 | (5) |
| AR | 8/11/20 | 8/25/20 | 3 | 1.76 | 4.35 | (5) |
| AZ | 8/12/20 | 8/26/20 | 4.7 | 2.7 | 7.61 | (5) |
| CA | 8/13/20 | 8/19/20 | 4.3 | 2.85 | 6.06 | (5) |
| CO | 8/10/20 | 8/25/20 | 3.3 | 1.83 | 4.8 | (5) |
| CT | 8/11/20 | 8/24/20 | 2.4 | 1.45 | 3.52 | (5) |
| DC | 8/13/20 | 8/27/20 | 6.8 | 4.59 | 9.21 | (5) |
| DE | 8/12/20 | 8/27/20 | 8.5 | 5.79 | 11.25 | (5) |
| FL | 8/14/20 | 8/14/20 | 4.5 | 3.15 | 6.06 | (5) |
| GA | 8/13/20 | 8/26/20 | 7.4 | 5.52 | 9.51 | (5) |
| HI | 8/14/20 | 8/26/20 |  |  |  | (5) |
| IA | 8/12/20 | 8/27/20 | 9.4 | 7.12 | 11.63 | (5) |
| ID | 8/12/20 | 8/26/20 | 4.6 | 1.67 | 9.05 | (5) |
| IL | 8/12/20 | 8/27/20 | 4.9 | 3.4 | 6.65 | (5) |
| IN | 8/12/20 | 8/26/20 | 3.1 | 1.53 | 4.76 | (5) |
| KS | 8/11/20 | 8/25/20 | 3.7 | 2.02 | 5.47 | (5) |
| KY | 8/12/20 | 8/26/20 | 3.1 | 2.05 | 4.49 | (5) |
| LA | 8/12/20 | 8/25/20 | 11.8 | 9.27 | 14.81 | (5) |
| MA | 8/12/20 | 8/27/20 | 3.7 | 2.27 | 5.21 | (5) |
| MD | 8/10/20 | 8/26/20 | 7 | 4.92 | 9.33 | (5) |
| ME | 8/12/20 | 8/27/20 | 0.6 | 0 | 1.44 | (5) |
| MI | 8/12/20 | 8/25/20 | 4.2 | 2.88 | 5.76 | (5) |
| MN | 8/11/20 | 8/27/20 | 4.5 | 3.05 | 5.87 | (5) |
| MO | 8/12/20 | 8/21/20 | 3.5 | 2.23 | 5.06 | (5) |
| MS | 8/12/20 | 8/27/20 | 10 | 6.69 | 14.71 | (5) |
| MT | 8/12/20 | 8/24/20 | 1.3 | 0.38 | 2.49 | (5) |
| NC | 8/11/20 | 8/27/20 | 3.8 | 2.33 | 5.29 | (5) |
| ND | 8/12/20 | 8/26/20 | 0.6 | 0 | 1.45 | (5) |
| NE | 8/11/20 | 8/25/20 | 7.9 | 5.51 | 10.61 | (5) |
| NH | 8/13/20 | 8/25/20 | 0.8 | 0.3 | 1.22 | (5) |
| NJ | 8/10/20 | 8/26/20 | 12.2 | 10.15 | 14.53 | (5) |
| NM | 8/10/20 | 8/27/20 | 2.5 | 1.47 | 3.79 | (5) |
| NV | 8/12/20 | 8/26/20 | 7.9 | 6.13 | 9.83 | (5) |
| NY | 8/10/20 | 8/26/20 | 20.6 | 18.04 | 23.14 | (5) |
| OH | 8/12/20 | 8/27/20 | 2.1 | 1.06 | 3.29 | (5) |
| OK | 8/10/20 | 8/18/20 | 4 | 2.72 | 5.51 | (5) |
| OR | 8/10/20 | 8/27/20 | 2.4 | 1.41 | 3.6 | (5) |
| PA | 8/10/20 | 8/26/20 | 10.1 | 7.53 | 13.74 | (5) |
| PR | 8/10/20 | 8/17/20 | 2.2 | 1.18 | 3.26 | (5) |
| RI | 8/12/20 | 8/27/20 | 3.6 | 1.69 | 5.89 | (5) |
| SC | 8/12/20 | 8/27/20 | 6 | 4.37 | 7.95 | (5) |
| SD | 8/12/20 | 8/26/20 | NA | 0 | 4.35 | (5) |
| TN | 8/12/20 | 8/26/20 | 6.7 | 5.11 | 8.57 | (5) |
| TX | 8/12/20 | 8/24/20 | 6.5 | 4.69 | 8.47 | (5) |
| UT | 8/15/20 | 8/25/20 | 5.5 | 2.94 | 8.71 | (5) |
| VA | 8/10/20 | 8/26/20 | 4.7 | 2.51 | 7.22 | (5) |
| VT | 8/13/20 | 8/27/20 | 0.3 | 0 | 0.81 | (5) |
| WA | 8/12/20 | 8/27/20 | 3 | 1.79 | 4.47 | (5) |
| WI | 8/12/20 | 8/27/20 | 3.3 | 2.03 | 4.72 | (5) |
| WV | 8/13/20 | 8/27/20 | 2.2 | 0.52 | 4.9 | (5) |
| WY | 8/13/20 | 8/24/20 | 0.8 | 0 | 3.11 | (5) |
| AK | 8/26/20 | 9/9/20 | 1 | 0.24 | 2.17 | (5) |
| AL | 8/26/20 | 9/8/20 | 9.9 | 7.19 | 12.79 | (5) |
| AR | 8/24/20 | 9/8/20 | 4.9 | 3.5 | 6.41 | (5) |
| AZ | 8/26/20 | 9/9/20 | 4.9 | 3.05 | 7.11 | (5) |
| CA | 8/28/20 | 9/9/20 | 6 | 4.2 | 7.82 | (5) |
| CO | 8/24/20 | 9/4/20 | 3.6 | 2.31 | 5.09 | (5) |
| CT | 8/26/20 | 9/4/20 | 4.3 | 2.9 | 5.69 | (5) |
| DC | 8/27/20 | 9/10/20 | 5 | 2.83 | 7.55 | (5) |
| DE | 8/26/20 | 9/10/20 | 4.5 | 2.42 | 7.12 | (5) |
| FL | 8/27/20 | 9/9/20 | 5.7 | 3.93 | 7.49 | (5) |
| GA | 8/25/20 | 9/10/20 | 8.7 | 6.69 | 11.14 | (5) |
| HI | 8/28/20 | 9/9/20 |  |  |  | (5) |
| IA | 8/25/20 | 9/10/20 | 8.4 | 6.15 | 10.74 | (5) |
| ID | 8/26/20 | 9/10/20 | 5.7 | 2.9 | 9.7 | (5) |
| IL | 8/26/20 | 9/3/20 | 5.6 | 3.9 | 7.67 | (5) |
| IN | 8/26/20 | 9/9/20 | 2.4 | 1.36 | 3.52 | (5) |
| KS | 8/24/20 | 9/3/20 | 2.9 | 1.57 | 4.31 | (5) |
| KY | 8/26/20 | 9/8/20 | 3.1 | 1.91 | 4.33 | (5) |
| LA | 8/26/20 | 9/8/20 | 8.6 | 6.65 | 11.3 | (5) |
| MA | 8/27/20 | 9/5/20 | 3.6 | 2.36 | 5.08 | (5) |
| MD | 8/26/20 | 9/8/20 | 7.9 | 5.72 | 10.35 | (5) |
| ME | 8/26/20 | 9/10/20 | 1.5 | 0.44 | 3.22 | (5) |
| MI | 8/26/20 | 9/3/20 | 3.4 | 2.16 | 4.95 | (5) |
| MN | 8/26/20 | 9/8/20 | 9.2 | 6.71 | 11.95 | (5) |
| MO | 8/24/20 | 9/10/20 | 2.9 | 1.74 | 4.17 | (5) |
| MS | 8/26/20 | 9/10/20 | 8.4 | 4.89 | 12.73 | (5) |
| MT | 8/24/20 | 9/9/20 | 0.9 | 0.23 | 1.82 | (5) |
| NC | 8/26/20 | 9/9/20 | 3.8 | 2.33 | 5.26 | (5) |
| ND | 8/26/20 | 9/9/20 | 0.2 | 0 | 0.99 | (5) |
| NE | 8/24/20 | 9/10/20 | 6.3 | 4.47 | 8.27 | (5) |
| NH | 8/26/20 | 9/3/20 | 1.6 | 0.73 | 2.84 | (5) |
| NJ | 8/26/20 | 9/7/20 | 12.8 | 10.49 | 15.35 | (5) |
| NM | 8/25/20 | 9/8/20 | 3.7 | 2.28 | 5.55 | (5) |
| NV | 8/27/20 | 8/29/20 | 6.5 | 4.63 | 8.58 | (5) |
| NY | 8/26/20 | 9/10/20 | 19.5 | 16.88 | 22.38 | (5) |
| OH | 8/27/20 | 9/10/20 | 5 | 3.17 | 7.14 | (5) |
| OK | 8/24/20 | 9/1/20 | 5.4 | 3.83 | 7.29 | (5) |
| OR | 8/26/20 | 9/10/20 | 1.9 | 0.92 | 2.94 | (5) |
| PA | 8/26/20 | 9/9/20 | 9.5 | 6.58 | 13.56 | (5) |
| PR | 8/24/20 | 9/3/20 | 2.5 | 1.36 | 3.76 | (5) |
| RI | 8/27/20 | 9/10/20 | 3.1 | 1.73 | 4.64 | (5) |
| SC | 8/26/20 | 9/10/20 | 7.2 | 5.12 | 9.57 | (5) |
| SD | 8/26/20 | 9/10/20 | 0.7 | 0 | 2.47 | (5) |
| TN | 8/25/20 | 9/8/20 | 5.4 | 3.62 | 7.31 | (5) |
| TX | 8/25/20 | 9/2/20 | 5.8 | 4.04 | 7.92 | (5) |
| UT | 8/24/20 | 9/8/20 | 4.9 | 2.82 | 7.67 | (5) |
| VA | 8/26/20 | 9/8/20 | 3.5 | 1.97 | 5.26 | (5) |
| VT | 8/26/20 | 9/10/20 | 0.4 | 0 | 0.94 | (5) |
| WA | 8/25/20 | 9/10/20 | 4.5 | 2.63 | 6.94 | (5) |
| WI | 8/25/20 | 9/10/20 | 2.6 | 1.48 | 3.95 | (5) |
| WV | 8/26/20 | 9/10/20 | 1.5 | 0.78 | 2.38 | (5) |
| WY | 8/24/20 | 9/9/20 |  |  |  | (5) |
| AK | 9/9/20 | 9/18/20 | 0.4 | 0.11 | 0.82 | (5) |
| AL | 9/9/20 | 9/18/20 | 8.7 | 6.19 | 11.31 | (5) |
| AR | 9/9/20 | 9/18/20 | 6.3 | 4.6 | 8.03 | (5) |
| AZ | 9/9/20 | 9/23/20 | 5.4 | 3.08 | 8.52 | (5) |
| CA | 9/10/20 | 9/16/20 | 4.9 | 3.17 | 6.85 | (5) |
| CO | 9/9/20 | 9/18/20 | 3.3 | 1.9 | 4.92 | (5) |
| CT | 9/9/20 | 9/14/20 | 3.1 | 2.06 | 4.37 | (5) |
| DC | 9/8/20 | 9/24/20 | 6.5 | 4.47 | 8.29 | (5) |
| DE | 9/9/20 | 9/24/20 | 7.5 | 4.89 | 10.5 | (5) |
| FL | 9/11/20 | 9/11/20 | 8.5 | 6.55 | 10.68 | (5) |
| GA | 9/9/20 | 9/18/20 | 13 | 10.51 | 15.79 | (5) |
| HI | 9/8/20 | 9/22/20 | 0.8 | 0 | 2.18 | (5) |
| IA | 9/9/20 | 9/24/20 | 7.6 | 5.56 | 10.02 | (5) |
| ID | 9/8/20 | 9/18/20 | 5.2 | 2.74 | 8.46 | (5) |
| IL | 9/8/20 | 9/17/20 | 4.5 | 3.1 | 6.07 | (5) |
| IN | 9/9/20 | 9/23/20 | 4 | 1.44 | 8.79 | (5) |
| KS | 9/9/20 | 9/19/20 | 3.5 | 2.31 | 4.84 | (5) |
| KY | 9/9/20 | 9/18/20 | 3.6 | 2.33 | 5.21 | (5) |
| LA | 9/9/20 | 9/19/20 | 12.5 | 10.02 | 15.61 | (5) |
| MA | 9/9/20 | 9/15/20 | 3.7 | 2.23 | 5.16 | (5) |
| MD | 9/10/20 | 9/23/20 | 10.2 | 7.91 | 12.68 | (5) |
| ME | 9/8/20 | 9/24/20 | 0.5 | 0.09 | 0.87 | (5) |
| MI | 9/8/20 | 9/22/20 | 3.7 | 2.54 | 4.93 | (5) |
| MN | 9/9/20 | 9/23/20 | 8 | 4.74 | 11.43 | (5) |
| MO | 9/8/20 | 9/24/20 | 3.5 | 2.15 | 5.01 | (5) |
| MS | 9/9/20 | 9/22/20 | 7.9 | 5.35 | 10.87 | (5) |
| MT | 9/9/20 | 9/23/20 | 2.2 | 0.67 | 4.38 | (5) |
| NC | 9/9/20 | 9/17/20 | 6.8 | 4.83 | 8.87 | (5) |
| ND | 9/9/20 | 9/24/20 | 1.2 | 0 | 3.4 | (5) |
| NE | 9/9/20 | 9/18/20 | 6.7 | 4.8 | 8.88 | (5) |
| NH | 9/9/20 | 9/17/20 | 0.7 | 0.26 | 1.32 | (5) |
| NJ | 9/9/20 | 9/23/20 | 15.1 | 12.65 | 17.63 | (5) |
| NM | 9/8/20 | 9/24/20 | 2.4 | 1.19 | 3.65 | (5) |
| NV | 9/9/20 | 9/12/20 | 7.8 | 5.9 | 9.99 | (5) |
| NY | 9/11/20 | 9/24/20 | 17 | 14.72 | 19.23 | (5) |
| OH | 9/10/20 | 9/22/20 | 2.8 | 1.69 | 4.16 | (5) |
| OK | 9/9/20 | 9/18/20 | 5 | 3.44 | 6.54 | (5) |
| OR | 9/8/20 | 9/22/20 | 2.6 | 1.51 | 4.06 | (5) |
| PA | 9/9/20 | 9/23/20 | 11.1 | 8.85 | 13.71 | (5) |
| PR | 9/8/20 | 9/15/20 | 3 | 1.78 | 4.27 | (5) |
| RI | 9/9/20 | 9/24/20 | 2.7 | 1.15 | 4.91 | (5) |
| SC | 9/9/20 | 9/18/20 | 7.8 | 5.58 | 10.19 | (5) |
| SD | 9/9/20 | 9/23/20 | 1.8 | 0 | 4.82 | (5) |
| TN | 9/8/20 | 9/16/20 | 6.7 | 4.96 | 8.57 | (5) |
| TX | 9/9/20 | 9/24/20 | 8.2 | 6.16 | 10.47 | (5) |
| UT | 9/9/20 | 9/18/20 | 5.1 | 3.29 | 7.9 | (5) |
| VA | 9/10/20 | 9/23/20 | 3.2 | 1.81 | 5.13 | (5) |
| VT | 9/9/20 | 9/24/20 | 1.7 | 0.27 | 4.15 | (5) |
| WA | 9/9/20 | 9/22/20 | 2.5 | 1.29 | 3.83 | (5) |
| WI | 9/9/20 | 9/24/20 | 3.8 | 2.2 | 5.73 | (5) |
| WV | 9/9/20 | 9/22/20 | 1.3 | 0.51 | 2.18 | (5) |
| WY | 9/9/20 | 9/24/20 | 1.5 | 0 | 3.97 | (5) |

**Table S2:** Posterior distributions and convergence diagnostic of *n* and SP_o_ for individual states (random effects)

| **State** | **n: median [95% CrI]** | **n: PSRF** | **SPo/N (%): median [95% CrI]** | **SPo/N (%): PSRF** |
| --- | --- | --- | --- | --- |
| AL | 0.44 [0.37 - 0.56] | 1.05 | 0.58 [0 - 4.13] | 1.01 |
| AK | 0.5 [0.41 - 0.64] | 1.04 | 0.14 [0 - 0.59] | 1.01 |
| AZ | 0.49 [0.42 - 0.6] | 1.05 | 0.36 [0 - 2.59] | 1.01 |
| AR | 0.47 [0.4 - 0.59] | 1.05 | 0.31 [0 - 1.93] | 1.01 |
| CA | 0.43 [0.35 - 0.58] | 1.04 | 0.87 [0.01 - 3.39] | 1.02 |
| CO | 0.49 [0.42 - 0.6] | 1.04 | 0.3 [0 - 1.87] | 1.01 |
| CT | 0.47 [0.4 - 0.61] | 1.05 | 0.72 [0 - 3.01] | 1.03 |
| DE | 0.44 [0.37 - 0.57] | 1.04 | 0.64 [0 - 4.09] | 1.01 |
| DC | 0.44 [0.36 - 0.58] | 1.05 | 0.87 [0 - 4.5] | 1.01 |
| FL | 0.47 [0.39 - 0.59] | 1.06 | 0.46 [0 - 2.74] | 1.01 |
| GA | 0.43 [0.36 - 0.55] | 1.05 | 0.58 [0 - 4.16] | 1.01 |
| HI | 0.51 [0.4 - 0.64] | 1.03 | 0.13 [0 - 0.82] | 1.01 |
| ID | 0.46 [0.38 - 0.59] | 1.04 | 0.45 [0 - 3.08] | 1.01 |
| IL | 0.44 [0.36 - 0.6] | 1.04 | 1.43 [0.01 - 4.84] | 1.01 |
| IN | 0.51 [0.42 - 0.65] | 1.03 | 0.94 [0.02 - 2.19] | 1.03 |
| IA | 0.43 [0.35 - 0.56] | 1.05 | 0.94 [0.01 - 5.21] | 1.01 |
| KS | 0.51 [0.43 - 0.61] | 1.05 | 0.24 [0 - 1.45] | 1.01 |
| KY | 0.46 [0.38 - 0.59] | 1.04 | 0.55 [0 - 2.23] | 1.01 |
| LA | 0.45 [0.34 - 0.59] | 1.02 | 5.01 [1.63 - 8.57] | 1.01 |
| ME | 0.53 [0.45 - 0.65] | 1.03 | 0.11 [0 - 0.51] | 1.02 |
| MD | 0.42 [0.33 - 0.57] | 1.04 | 2.34 [0.01 - 7.54] | 1.01 |
| MA | 0.48 [0.4 - 0.6] | 1.04 | 0.39 [0 - 2.61] | 1.01 |
| MI | 0.46 [0.39 - 0.59] | 1.04 | 0.46 [0 - 2.6] | 1.02 |
| MN | 0.4 [0.32 - 0.55] | 1.05 | 0.96 [0.01 - 3.11] | 1.03 |
| MS | 0.45 [0.37 - 0.57] | 1.05 | 0.58 [0 - 4.29] | 1 |
| MO | 0.52 [0.42 - 0.64] | 1.04 | 1.19 [0.38 - 1.98] | 1.04 |
| MT | 0.49 [0.4 - 0.62] | 1.04 | 0.18 [0 - 0.87] | 1.03 |
| NE | 0.43 [0.36 - 0.57] | 1.05 | 0.74 [0 - 4.23] | 1.02 |
| NV | 0.44 [0.36 - 0.57] | 1.05 | 0.61 [0 - 4.04] | 1.01 |
| NH | 0.54 [0.46 - 0.65] | 1.04 | 0.12 [0 - 0.59] | 1.01 |
| NJ | 0.4 [0.32 - 0.57] | 1.05 | 2.06 [0.01 - 10.22] | 1.02 |
| NM | 0.46 [0.36 - 0.61] | 1.04 | 0.75 [0.01 - 2.3] | 1.01 |
| NY | 0.44 [0.32 - 0.59] | 1.02 | 14.19 [7.54 - 20.16] | 1 |
| NC | 0.46 [0.38 - 0.58] | 1.05 | 0.36 [0 - 2.18] | 1.01 |
| ND | 0.52 [0.42 - 0.66] | 1.03 | 0.32 [0 - 1.57] | 1.03 |
| OH | 0.47 [0.4 - 0.6] | 1.05 | 0.38 [0 - 1.9] | 1.01 |
| OK | 0.45 [0.38 - 0.57] | 1.06 | 0.31 [0 - 1.9] | 1.01 |
| OR | 0.46 [0.37 - 0.61] | 1.04 | 0.69 [0.01 - 2.02] | 1.02 |
| PA | 0.41 [0.31 - 0.58] | 1.03 | 3.36 [0.02 - 8.3] | 1.01 |
| RI | 0.51 [0.43 - 0.63] | 1.05 | 0.24 [0 - 1.57] | 1.02 |
| SC | 0.44 [0.37 - 0.56] | 1.05 | 0.46 [0 - 3.34] | 1.02 |
| SD | 0.54 [0.44 - 0.67] | 1.03 | 0.19 [0 - 1.34] | 1.01 |
| TN | 0.43 [0.35 - 0.56] | 1.04 | 0.56 [0 - 3.15] | 1.01 |
| TX | 0.44 [0.37 - 0.57] | 1.05 | 0.57 [0 - 3.17] | 1.02 |
| UT | 0.47 [0.39 - 0.59] | 1.04 | 0.93 [0.21 - 1.88] | 1 |
| VT | 0.51 [0.42 - 0.64] | 1.04 | 0.13 [0 - 0.56] | 1.01 |
| VA | 0.46 [0.38 - 0.6] | 1.04 | 0.91 [0.01 - 3.54] | 1.02 |
| WA | 0.45 [0.37 - 0.6] | 1.03 | 0.82 [0.01 - 2.65] | 1.01 |
| WV | 0.47 [0.39 - 0.61] | 1.04 | 0.31 [0 - 1.18] | 1.01 |
| WI | 0.49 [0.41 - 0.61] | 1.04 | 0.3 [0 - 1.7] | 1.01 |
| WY | 0.5 [0.4 - 0.63] | 1.03 | 0.22 [0 - 1.41] | 1.02 |

**Table S3.** Posterior estimates of prevalence and seroprevalence as of December 8, 2020.

| **state** | **Prevalence median [95% CrI]** | **Prevalence 2-wk change in median** | **Seroprevalence median [95% CrI]** |
| --- | --- | --- | --- |
| AK | 0.7% [0.41%-1.07%] | +0.05% | 3.5% [2.3%-4.8%] |
| AL | 2.23% [1.11%-3.5%] | +0.62% | 15.3% [11.1%-20.4%] |
| AR | 1.26% [0.67%-1.86%] | +0.22% | 9.4% [7%-12%] |
| AZ | 1.76% [0.88%-2.69%] | +0.59% | 9.9% [7.1%-13.2%] |
| CA | 1.23% [0.58%-1.84%] | +0.56% | 7.8% [6%-10.3%] |
| CO | 1.49% [0.77%-2.29%] | -0.01% | 8.6% [6%-11.6%] |
| CT | 0.81% [0.44%-1.11%] | +0.23% | 6.8% [5.6%-8.3%] |
| DC | 0.51% [0.27%-0.75%] | +0.19% | 7.9% [6%-10.4%] |
| DE | 1.97% [0.93%-2.95%] | +0.71% | 10.9% [8.2%-14.5%] |
| FL | 1.15% [0.55%-1.83%] | +0.18% | 10.7% [7.9%-14.2%] |
| GA | 1.11% [0.56%-1.66%] | +0.42% | 12% [9.3%-15.5%] |
| HI | 0.83% [0.24%-2.2%] | +0.06% | 4.8% [2%-9.7%] |
| IA | 2.16% [0.87%-3.59%] | -0.65% | 20.3% [13.7%-27.5%] |
| ID | 2.51% [1.14%-4.05%] | +0.27% | 14.9% [9.8%-21.2%] |
| IL | 1.08% [0.51%-1.66%] | -0.14% | 11.5% [8.5%-14.9%] |
| IN | 1.82% [0.79%-3.06%] | +0.23% | 9.8% [6%-14.1%] |
| KS | 1.72% [0.92%-2.77%] | -0.03% | 10.4% [7.2%-14.4%] |
| KY | 1.2% [0.61%-1.84%] | +0.14% | 7.4% [5.3%-9.7%] |
| LA | 0.91% [0.44%-1.59%] | +0.12% | 14.6% [11.3%-18.9%] |
| MA | 1.39% [0.71%-2.17%] | +0.62% | 7.7% [5.8%-10.2%] |
| MD | 1.36% [0.55%-2.28%] | +0.2% | 13.4% [10.2%-17.9%] |
| ME | 0.28% [0.16%-0.42%] | +0.13% | 1.4% [1%-2%] |
| MI | 0.97% [0.49%-1.42%] | -0.12% | 7.7% [5.8%-10%] |
| MN | 2.44% [1.07%-3.8%] | -0.3% | 15.9% [10.7%-21%] |
| MO | 1.35% [0.6%-2.42%] | -0.07% | 9.4% [6.2%-13.6%] |
| MS | 1.73% [0.83%-2.67%] | +0.42% | 14.3% [10.6%-19.2%] |
| MT | 1.12% [0.57%-1.8%] | -0.26% | 7.5% [4.8%-10.6%] |
| NC | 0.95% [0.53%-1.42%] | +0.3% | 7.3% [5.5%-9.8%] |
| ND | 1.65% [0.66%-3.07%] | -0.91% | 15% [8.2%-23%] |
| NE | 2.61% [1.11%-4.1%] | -0.31% | 17.9% [12.3%-24.4%] |
| NH | 0.92% [0.47%-1.48%] | +0.29% | 2.8% [1.9%-4.1%] |
| NJ | 1.14% [0.49%-1.7%] | +0.14% | 15.9% [12.4%-20.2%] |
| NM | 1.28% [0.58%-2.09%] | -0.45% | 7.7% [5.4%-10.5%] |
| NV | 2.69% [1.19%-4.34%] | +0.46% | 15.1% [10.6%-20.3%] |
| NY | 0.66% [0.36%-1.22%] | +0.23% | 22.3% [17.9%-27.5%] |
| OH | 1.51% [0.78%-2.2%] | +0.32% | 7% [5.1%-9.4%] |
| OK | 1.42% [0.75%-2.12%] | +0.06% | 9.7% [6.9%-12.8%] |
| OR | 0.54% [0.26%-0.85%] | +0.27% | 5.1% [3.5%-7.2%] |
| PA | 2.98% [1.09%-5.69%] | +0.93% | 15.8% [11%-22.2%] |
| RI | 1.81% [0.94%-2.83%] | +0.34% | 8.4% [5.7%-11.5%] |
| SC | 1.04% [0.55%-1.54%] | +0.41% | 10.4% [7.9%-13.4%] |
| SD | 1.57% [0.76%-3.04%] | -0.12% | 12% [6.3%-19.8%] |
| TN | 1.91% [0.98%-2.83%] | +0.5% | 11.7% [8.5%-15.4%] |
| TX | 1.23% [0.6%-1.84%] | +0.2% | 11.8% [8.9%-15.5%] |
| UT | 2% [0.99%-3.29%] | +0.1% | 12.7% [8.7%-17.3%] |
| VA | 0.84% [0.38%-1.34%] | +0.23% | 7.8% [5.6%-10.5%] |
| VT | 0.36% [0.16%-0.61%] | +0.11% | 1.6% [1%-2.5%] |
| WA | 0.71% [0.33%-1.13%] | -0.56% | 6.3% [4.5%-8.6%] |
| WI | 1.75% [0.78%-2.81%] | -0.2% | 12.1% [8%-16.5%] |
| WV | 0.89% [0.45%-1.35%] | +0.19% | 4.3% [2.9%-5.9%] |
| WY | 1.71% [0.8%-3.15%] | -0.44% | 9.8% [5.4%-16.1%] |
| **US** | **1.37% [0.76%-1.87%]** | **+0.24%** | **11.1% [10.1%-12.2%]** |

**Table S4.** International seroprevalence data.

| **Nation/ Country** | **Min date** | **Max date** | **Point est. (%)** | **LCL (%)** | **UCL (%)** | **Source** |
| --- | --- | --- | --- | --- | --- | --- |
| Belgium | 3/30/20 | 4/5/20 | 2.9 | 2.4 | 3.5 | (18) |
| Belgium | 4/20/20 | 4/26/20 | 6 | 5.2 | 6.9 | (18) |
| Belgium | 5/18/20 | 5/25/20 | 6.9 | 6.1 | 7.8 | (18) |
| Belgium | 6/8/20 | 6/13/20 | 5.5 | 4.7 | 6.4 | (18) |
| Belgium | 6/29/20 | 7/4/20 | 4.5 | 3.8 | 5.3 | (18) |
| Canada | 5/19/20 | 6/18/20 | 0.7 | 0.6 | 0.8 | (19) |
| Denmark | 4/6/20 | 5/3/20 | 1.9 | 0.8 | 2.3 | (20) |
| England | 2/1/20 | 3/31/20 | 1.8 | 0.8 | 3.5 | (21) |
| England | 2/1/20 | 3/31/20 | 1.4 | 0.1 | 5.9 | (21) |
| England | 3/16/20 | 6/30/20 | 3.9 | 3.4 | 4.4 | (22) |
| England | 4/1/20 | 4/30/20 | 4.5 | 2.1 | 8.4 | (21) |
| England | 4/1/20 | 4/30/20 | 6.6 | 4.8 | 8.8 | (21) |
| England | 4/26/20 | 7/26/20 | 6.2 | 5.6 | 6.9 | (23) |
| England | 4/26/20 | 7/8/20 | 6.3 | 5.6 | 7.1 | (24) |
| England | 4/30/20 | 5/22/20 | 8.6 | 8 | 9.2 | (25) |
| England | 5/1/20 | 5/31/20 | 2.7 | 0.7 | 0.9 | (21) |
| England | 5/1/20 | 5/31/20 | 7.8 | 6.3 | 9.6 | (21) |
| England | 5/22/20 | 6/8/20 | 8.2 | 7.5 | 8.9 | (25) |
| England | 6/1/20 | 8/2/20 | 3.2 | 2.5 | 4.1 | (21) |
| England | 6/1/20 | 8/2/20 | 4.9 | 2.7 | 8.1 | (21) |
| England | 6/8/20 | 7/6/20 | 7.1 | 6.6 | 7.6 | (25) |
| England | 6/20/20 | 7/13/20 | 6 | 5.8 | 6.1 | (26) |
| England | 7/13/20 | 7/21/20 | 6.8 | 6.3 | 7.3 | (25) |
| England | 7/20/20 | 8/16/20 | 5.5 | 5 | 6 | (25) |
| Greece | 3/1/20 | 3/31/20 | 0.02 | 0 | 0.25 | (27) |
| Greece | 4/1/20 | 4/30/20 | 0.25 | 0.02 | 0.5 | (27) |
| Hungary | 5/1/20 | 5/16/20 | 0.68 | 0.5 | 0.85 | (28) |
| Iceland | 4/1/20 | 4/4/20 | 0.6 | 0.3 | 0.9 | (29) |
| India | 5/11/20 | 5/25/20 | 0.7 | 0.6 | 0.8 | (9) |
| Ireland | 6/22/20 | 7/16/20 | 1.7 | 1.1 | 2.4 | (30) |
| Luxembourg | 4/15/20 | 5/5/20 | 2.06 | 1.34 | 2.77 | (31) |
| Norway | 4/20/20 | 5/17/20 | 1.1 | 0.5 | 2 | (32) |
| Portugal | 5/21/20 | 7/8/20 | 1.9 | 1.4 | 2.5 | (33) |
| Russia | 6/10/20 | 6/10/20 | 14 | 13.9 | 14.1 | (34) |
| Slovenia | 4/20/20 | 5/1/20 | 0.15 | 0.03 | 0.47 | (35) |

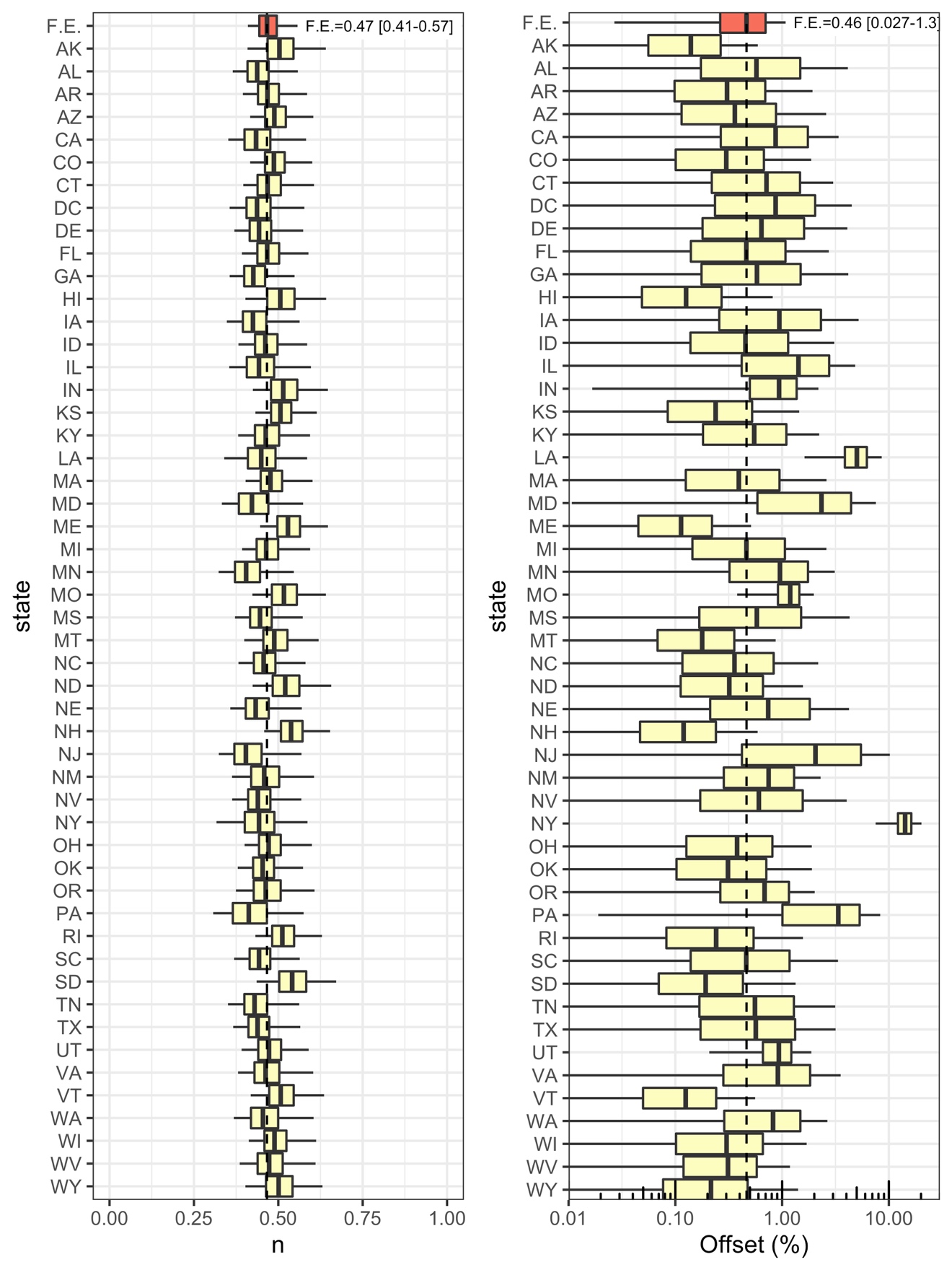

**Figure S1:** Posterior distributions of the power parameter n and the seroprevalence offset SP_o_ for individual states. The fixed effected is denoted by “F.E.,” and the vertical dashed line represents its posterior median.

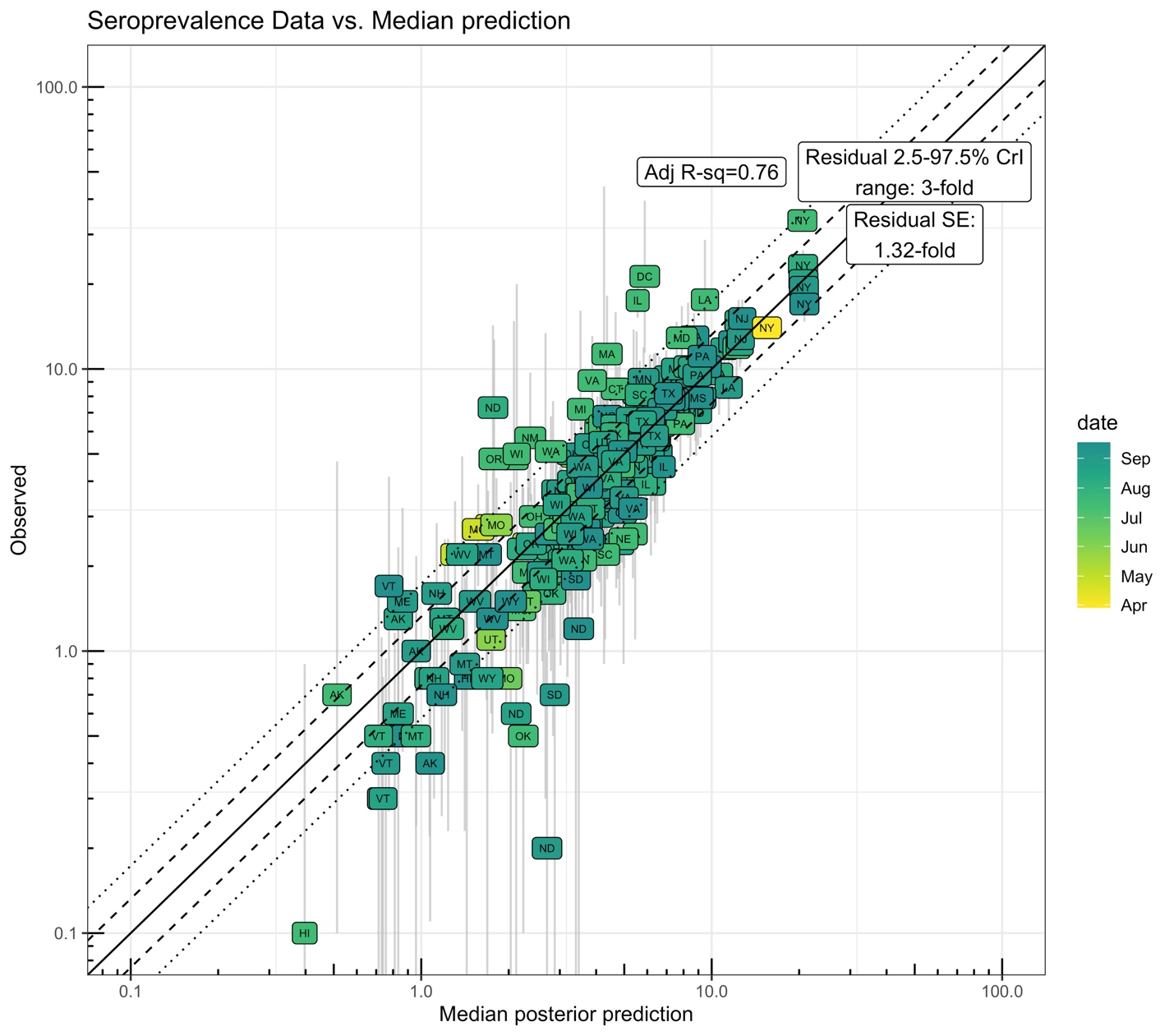

**Figure S2:** Scatter plot of seroprevalence predictions (posterior median) versus data (reported point estimate and 95% CI). The solid line represents equality, the dashed line is +/- one residual standard error, and the dotted line is the 95% CrI residual error. The adjusted R^2^ is calculated from a linear model based on the log-transformed posterior medians and the observed point estimates.

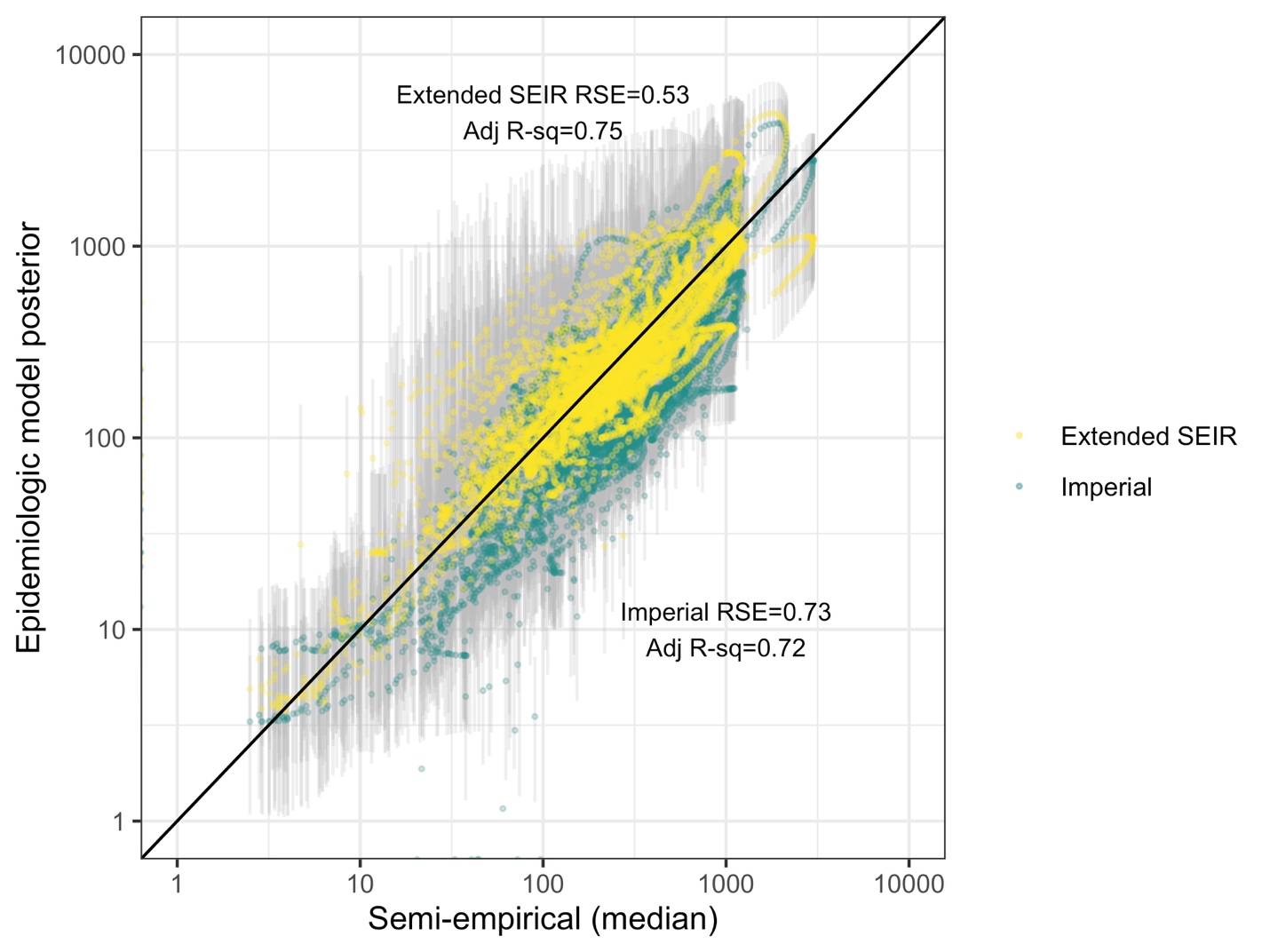

**Figure S3:** Scatter plot of active infection prevalence predictions from semi-empirical model (posterior median) versus those from epidemiologic models (posterior median and 95% CrI). The solid line represents equality. The residual standard error (RSE) and adjusted R^2^ are from the comparison of natural log-transformed median predictions.

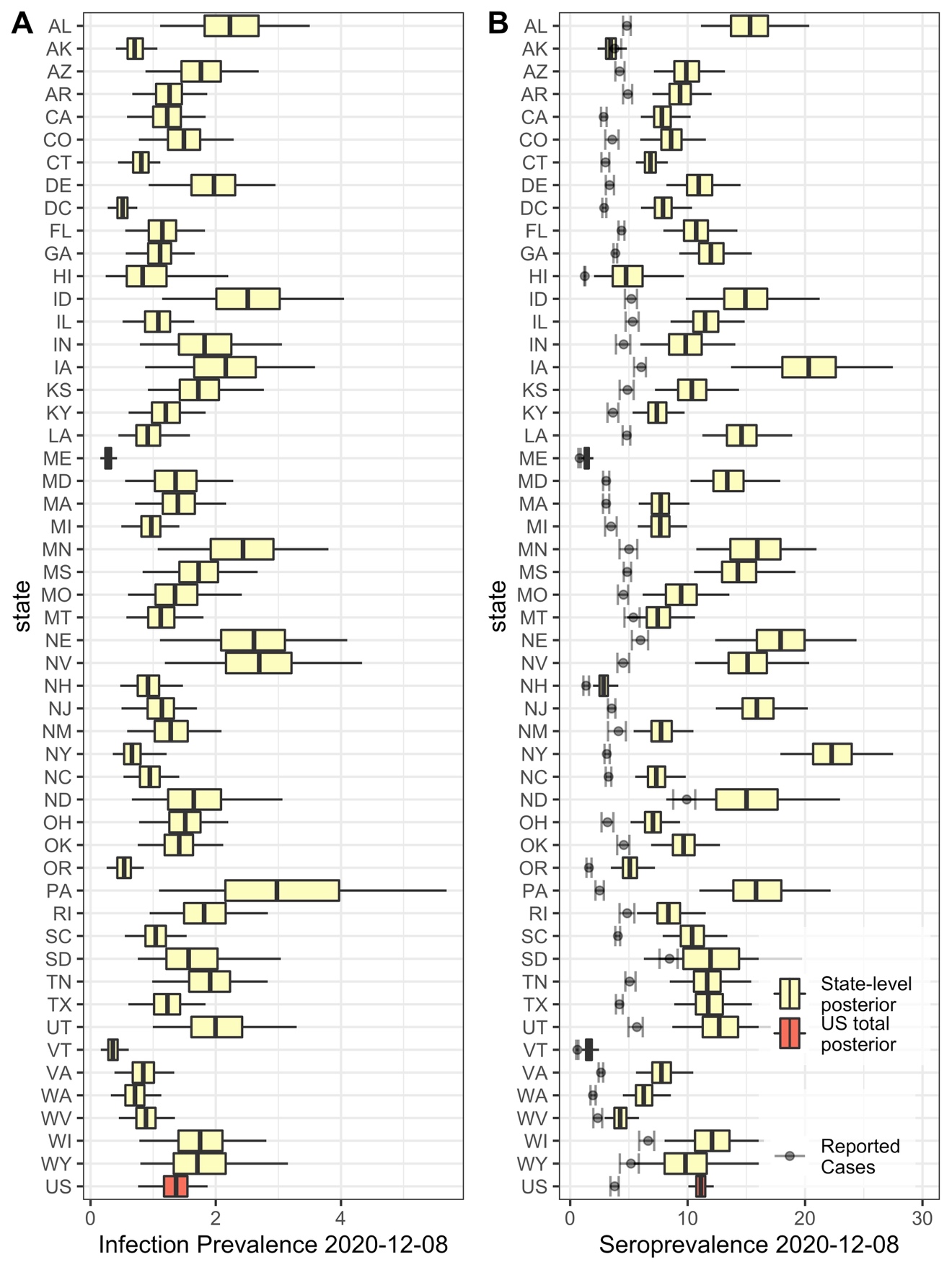

**Figure S4:** Boxplots (box=IQR, line=median, whiskers=95% CrI) of posterior estimate of infection prevalence (**A**) and seroprevalence (**B**) across states and for the U.S. overall as of December 8, 2020. In (B), for comparison, cumulative reported cases are shown with a 14-day lag to allow time for seroconversion (error bars denote range of 7-21 day lags).

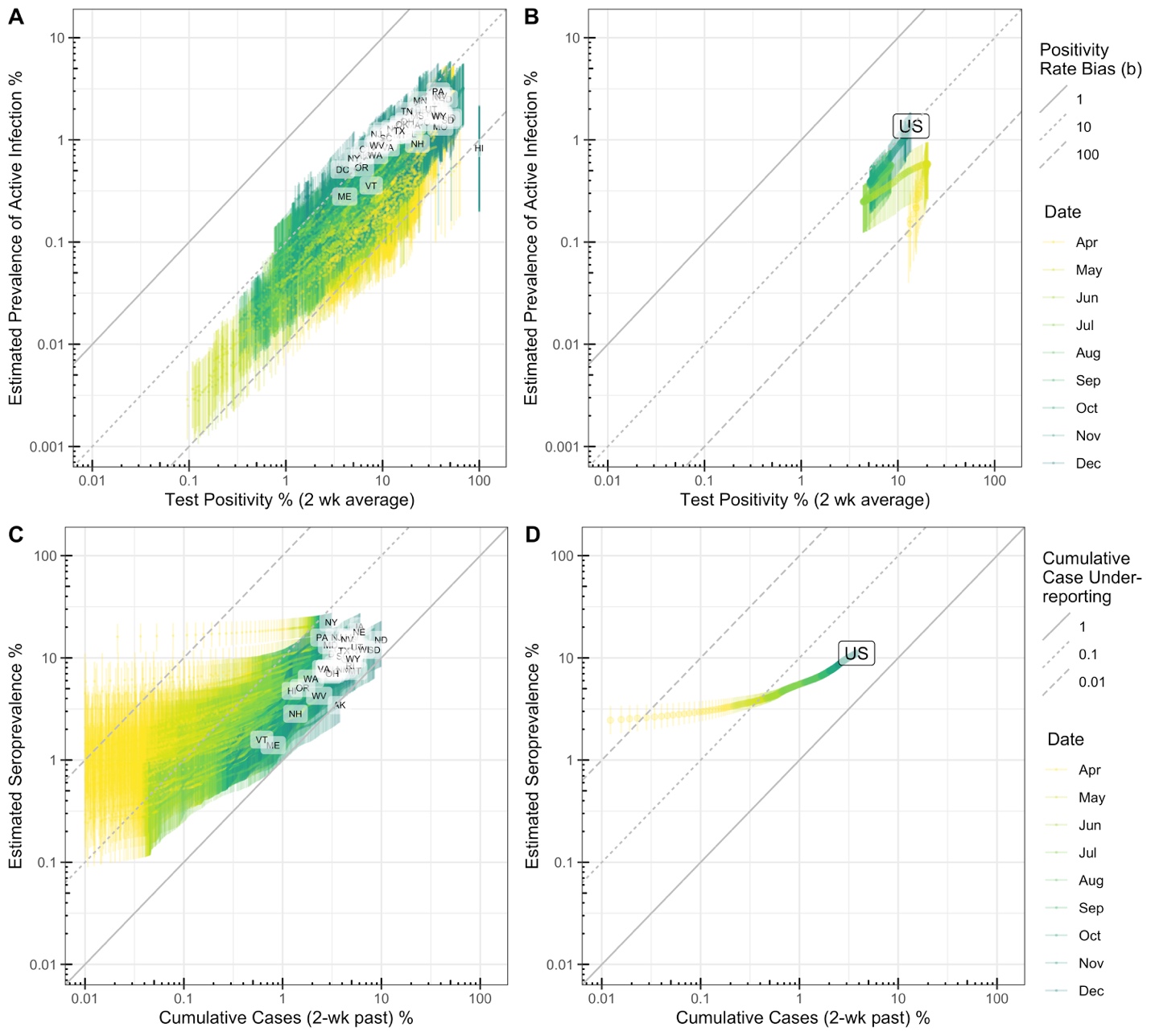

**Figure S5.** Bias estimates. **A,B)** Comparison of test positivity (14-day average) and semi-empirical prevalence estimates (median and 95% CrI) across all states (**A**) or across the U.S. in aggregate (**B**) from April 1-December 8, 2020. Diagonal lines denote different levels of positivity bias, as illustrated in Figure 1A. **C,D)** Comparison of cumulative reported cases, with 14-day lag to allow for conversion to seropositivity, and semi-empirical seropositivity estimates (median and 95% CrI) across all states (**C**) or across the U.S. in aggregate (**D**) from April 1-December 8, 2020. Diagonal lines denote different levels of cumulative case under-reporting.

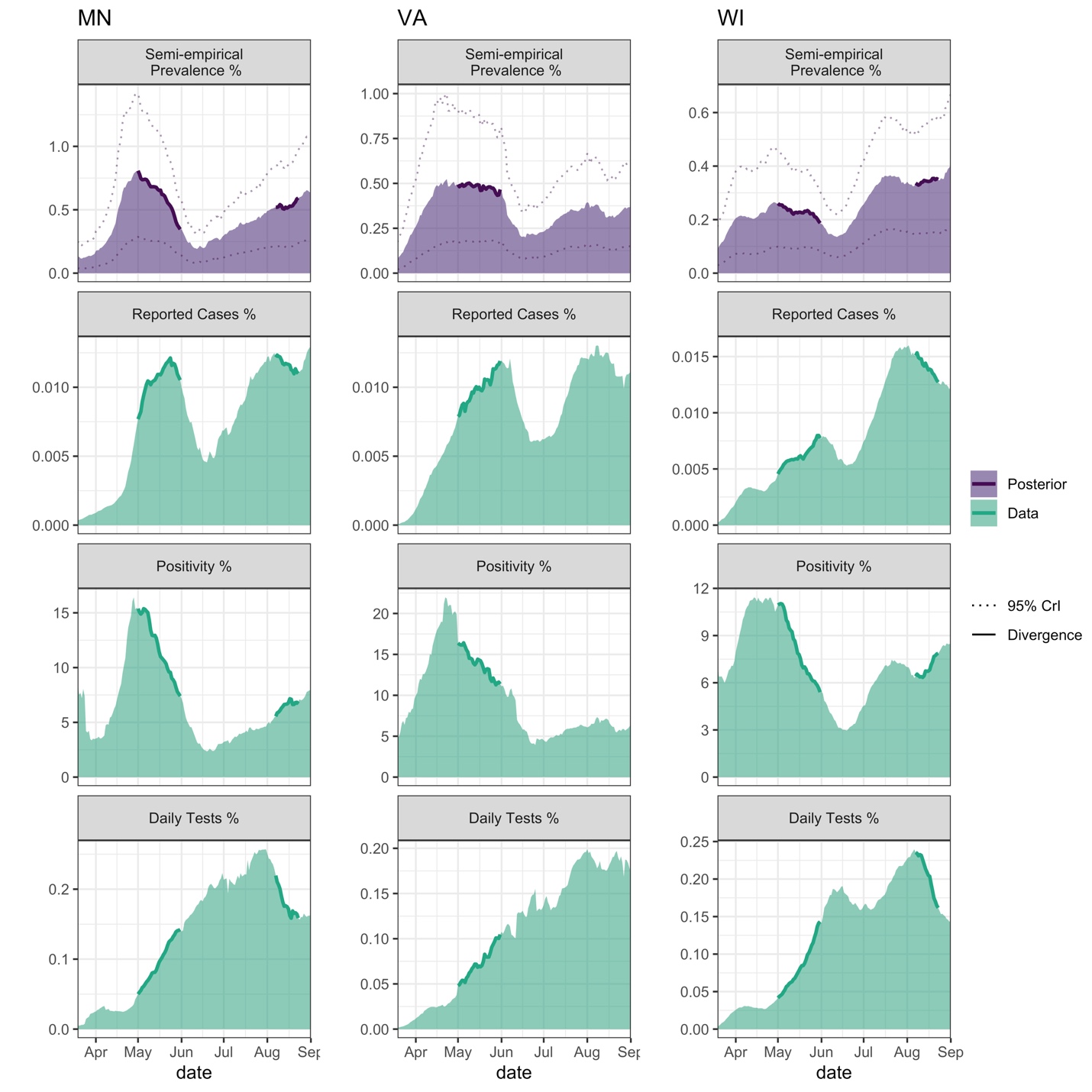

**Figure S6:** Examples of three states where in May or August, 2020, the trends in reported case rates and positivity rates diverged (i.e., one trending positive, the other negative). For each state, the top panel is the active infection prevalence as predicted by the semi-empirical model (posterior median and 95% CrI) whereas the bottom three panels show the reported case, positivity, and testing rates, each averaged over the previous 14 days. In May, reported cases were rising substantially in MN, VA, and WI at the same time that the test positivity rate was declining, testing rate was increasing, and the model predicted prevalence was flat or decreasing. By contrast, in August, the states of MN, VA, and WI all showed declining reported case rates while positivity was increasing. Our model predicts that COVID-19 prevalence was either flat or increasing during this time. In both scenarios, the increase (decrease) in reported cases was due to expanded (declining) testing rates, respectively, illustrating the pitfalls of relying on reported cases or test positivity rate alone to estimate the course of the epidemic.

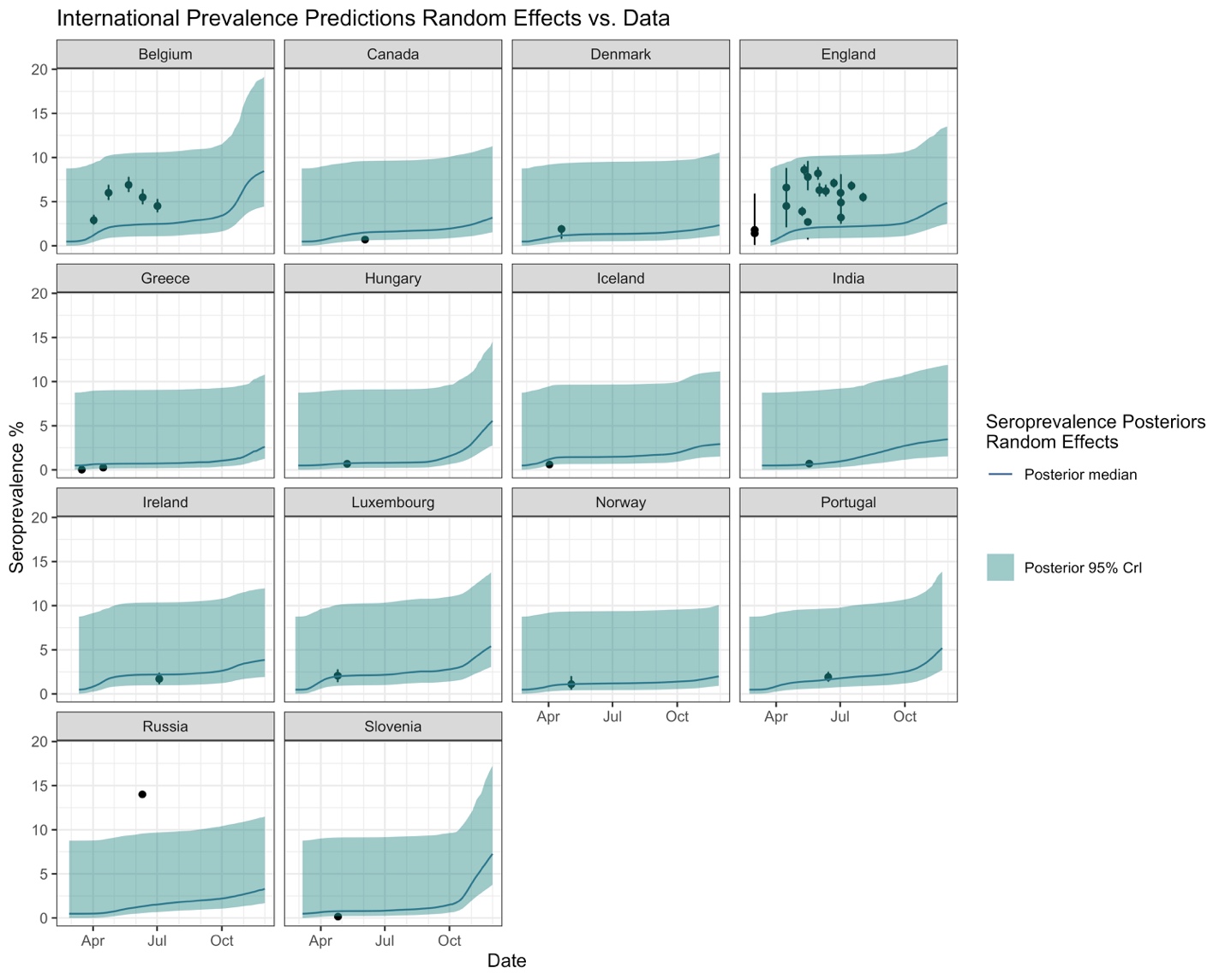

**Figure S7:** Application of semi-empirical model using random effects posterior distributions from U.S. states to other nations/countries. COVID-19 antibody seroprevalence estimates (posterior median and 95% credible intervals) for each nation/country with state-wide seroprevalence data (**Table S4**, reported point estimates and 95% confidence intervals shown).
